# Core Components for Emergency Medical Dispatch Systems: An International Delphi Consensus Study

**DOI:** 10.64898/2026.05.26.26354117

**Authors:** Katinka Weber, Willem Stassen, Sudha Jayaraman, Maria Lisa Odland, Aurore Nishimwe, Indrakantha Welgama, Lee Wallis, Agnieszka Ignatowicz, Justine Davies, Dispatch Guidance Working Group

## Abstract

**Introduction:** Emergency Medical Dispatch Systems (EMDS) can reduce delays in accessing emergency care by providing structured communication, triage, and coordination. However, such systems remain absent or underdeveloped in most low- or middle-income countries (LMICs). This study aimed to establish international consensus on essential EMDS components to inform global guidance.

**Methods:** We convened a multidisciplinary expert group to draft a preliminary list of essential components for three EMDS levels reflecting resource availability and system maturity. We then conducted a three-round Delphi with international experts to reach consensus on core EMDS components. Components which had ≥75% agreement were included, those with ≥75% disagreement were excluded. Components not achieving consensus by Round 3 were removed. Results were analysed overall and stratified by respondents’ country income level. A subsequent online expert meeting resolved inconsistencies and finalised the component list.

**Results:** The expert group generated 111 components for each of three EMDS levels (Foundational, Emerging, and Established) spanning 11 operational domains. Of the 68 experts invited to the Delphi, 43 participated in Round 1 and 30 in Round 3. Across all Delphi rounds, 289 components reached consensus for inclusion. The consensus resulted in a final list of 227 components (63 Foundational, 84 Emerging, and 80 Established). Consensus agreement clustered around core EMDS domains including communication, structured call-taking and prioritisation, advice-giving, resource dispatch and tracking, and foundational governance and data functions, whereas items showing either non-consensus or consensus disagreement were typically technology-dependent or context-specific.

**Conclusions:** This international consensus offers guidance for EMDS development across diverse resource settings and provides a scalable roadmap to strengthen emergency care systems.

## INTRODUCTION

Emergency conditions require urgent medical or surgical care to reduce the risk of death or long-term disability. These conditions account for an estimated 45-55% of global mortality, with the burden disproportionately affecting low- or middle-income countries (LMICs).^1–4^ It is estimated that around 40% of emergency deaths could be avoided if delays in accessing quality care were reduced.^5^ ^6^ Delays can be reduced by introducing prehospital emergency medical services, as well as ambulances, to assess, manage, and transport patients from the site of the emergency to the closest hospital able to treat them.^7^ ^8^ However, prehospital emergency medical services can also be associated with increased delays and death or disability.^9^ ^10^ At a minimum, good coordination between callers, ambulances, and health facilities is needed if the benefits of these systems are to be realised.^7^ ^8^

An efficient Emergency Medical Dispatch Systems (EMDS) provides this coordination, linking the caller, ambulance crews, health facilities, and, if required, other emergency services such as police or fire.^11^ Beyond basic communication and coordination, EMDS in well-resourced - often high-income country (HIC) settings - generally operate within established health system governance structures, including use of standardised guidance, protocols, trained personnel, and mechanisms for quality assurance and performance improvement.^8^ ^12^ Well-resourced EMDS also often utilise structured scripts and electronic decision-support tools to triage calls and, when multiple possible facilities are available to treat patients, destination triage can be used to direct patients to appropriate receiving facilities.^13^ In contrast, many lower resourced settings, as is the case in many LMICs, lack comprehensive EMDS infrastructure, relying instead on nascent systems or on imported models that might be ill-suited to local resources, leading to inefficiencies and poor patient outcomes.^1^ ^14^ ^15^

The transfer of HIC-based EMDS models to LMIC settings is constrained not only by funding availability, but also by contextual differences, for example in workforce capacity, demography and epidemiology, and health service availability.^15^ When transferred without contextual adaptation, these systems might fail or worsen efficiencies, which can cost lives.^1^ ^16^ The lack of adequate healthcare data in many LMICs risks that these issues are not detected.^17^ Even where bespoke systems are developed in LMICs, they are frequently implemented in isolation, with limited opportunities for knowledge sharing across countries.^17–19^

There is growing desire in many LMICs to strengthen their prehospital emergency medical services, and there is a critical gap in context-appropriate guidance and tools to support the establishment and development of EMDS for their coordination. The need to develop guidance and tools for establishing and developing EMDS for multiple contexts has global support, as expressed as a mandate in World Health Assembly (WHA) resolution 76.2 on Integrated Emergency, Critical and Operative Care for Universal Health Coverage and Protection from Health Emergencies.^20^ However, while EMDS principles are universally applicable, high-quality EMDS must align local contexts including burden of disease, geography, health service availability, population health literacy, infrastructure, local and regional cultural and political considerations, and available human and financial resources.^21–23^

This study aimed to achieve expert consensus on the key components of an EMDS system to enable development of essential tools and guidance for EMDS in a range of resource contexts. Such guidance is needed to inform the development of a scalable roadmap for countries or regions at different stages of EMDS development.

## METHODS

### Study Design

We utilised a modified Delphi methodology which provides a structured and iterative approach to achieving consensus among a diverse panel of experts across multiple contexts and countries.^24–26^ By engaging participants from a wide range of geographical, professional, and health system backgrounds, we aimed to develop globally relevant recommendations for components of EMDS that can be adapted to varying levels of resources and system maturity.

Our approach had three phases. First, during an in-person meeting in March 2025, a core team (JD, LW, AI, AN, WS, SJ, and VN) with experience in EMDS across LMICs conceptualised different levels of EMDS, based on resource availability and service delivery capacity. Then, guided by the WHO health system building blocks (health workforce, health information systems, access to essential medicines, health systems financing, leadership and governance, and service delivery)^27^ and experience of processes within EMDS, we developed an initial, hypothesised list of essential components across conceptual dispatch domains.

Second, between June and August 2025, we conducted a three-round modified Delphi process to develop consensus on the core components for each domain at each EMDS level using an electronic survey developed in REDCap (Research Electronic Data Capture, version 15.7.4, Vanderbilt University, Nashville, TN, USA).^28^ The survey was piloted among the study investigators prior to each round to ensure clarity and comprehension. In each round participants were asked to indicate agreement with the minimum essential components, processes, or tools (hereafter “components”) relevant to each of the domains and levels of EMDS, with agreement rated on a 5-point Likert scale ranging from strongly disagree to strongly agree, with a ‘not applicable’ option. In the first round, at the end of each domain, free-text fields were provided for participants to suggest additional components or comment on proposed items for each EMDS level. Additional components proposed in free-text from the first round were deduplicated and included in the second round. Components proceeded to subsequent rounds or were eliminated according to the consensus threshold. In Rounds 2 and 3, participants were presented with components from the preceding round which had not achieved agreement, along with information on the percentage agreement of respondents in the previous round. Participants were given two weeks to complete each round. Between rounds there was a one-week period for analysis and preparation of the subsequent round.

Demographic and professional information was collected, including age, sex, country of main employment, workplace type, job role in emergency medical dispatch, operational level of EMDS involvement (e.g. community, regional, national), years of experience working in EMDS, and formal medical or clinical training.

Third, we convened an online consensus meeting with selected Delphi participants to finalise the component list for each EMDS level. Participants for this meeting were sent final results from Round 3 prior to the meeting and were signposted to components where there was inconsistency in responses within and across levels (e.g., consensus on using a computer-aided dispatch system for ambulances alongside consensus on capturing all data on paper).

### Participants

Our aim was to gather responses from a diverse range of professionals with experience in EMDS across a broad range of geographical regions and country income strata, including clinical care providers, dispatchers, EMS leaders, policymakers, NGO representatives, and academics. We utilised our networks and snowball sampling to recruit participants. Participation was voluntary, with respondents given the option to continue into subsequent rounds. Knowledge of EMDS and consent to participate were eligibility criteria. Participants were invited to Rounds 2 or 3 if they had completed the previous round. Participants for the online consensus meeting were purposively selected from the participants of Round 3 of the Delphi survey, to allow for a maximum diversity of participants according to their characteristics and ensure that the number of participants was feasible to allow consensus to be developed in an online meeting.

### Data Analysis

Participant characteristics were summarised using counts and proportions. Delphi responses were calculated as the percentage of all respondents who *agreed* or *strongly agreed, or disagreed* or *strongly disagreed,* for inclusion of each component in each level of EMDS. Consensus to include in or exclude an EMDS component from the next round was indicated by achievement of a threshold of ≥75% *agreeing* or *strongly agreeing,* or *disagreeing* or *strongly disagreeing*, respectively.

The main analysis included all participants. Subgroup analyses on the responses from the final round were stratified by country income group (LMIC or HIC),^29^ based on participants’ primary country of work.

Component lists for each system level were derived using Round 3 results in conjunction with participants’ country income settings, based on the assumption that respondents are well positioned to assess feasibility within their respective resource contexts (with LMIC respondents’ responses used for Foundational components, HIC respondents for Established components, and all participants for Emerging components). Data were analysed in Microsoft Excel (for Microsoft 365 MSO, Version 2601 Build 16.0.19628.20214, 64-bit) to derive consensus outcomes.

### ETHICS

This study was reviewed by the University of Birmingham Research Ethics Committee and was determined to be low-risk with no full review being required (ERN_4117-Apr2025).

## RESULTS

### 1. Categorization across Foundational, Emerging, Established systems

The initial expert meeting agreed three broad EMDS levels applicable to countries with a spectrum of resource availability. A *Foundational EMDS* includes the essential components required for basic system functioning and coordination of EMDS. An *Emerging EMDS* builds on this foundation by integrating newer technologies and standards, thereby enhancing EMDS capabilities. An *Established EMDS* further advances this model through integration of more advanced technologies and protocols, enabling the most advanced EMDS. Experts also agreed 111 potential essential components for EMDS of any level, in 11 key structural or functional domains Table 1, below).

**Table 1.** 11 EMDS Key Domains.

| DOMAIN | DESCRIPTION |
| --- | --- |
| 1. Essential products and technologies | Hardware, software, and other essential products and technologies needed for the EMDS to work |
| 2. Call taking | Requirements for identification of the emergency and caller location |
| 3. Emergency prioritisation (triage) | Requirements for assessment of the urgency for resource (ambulance or other resource – e.g.: community first responder) dispatch to patients |
| 4. Advice giving | Guidance provided to the caller, from general instructions to life saving interventions |
| 5. Resource dispatch | Requirements for determining and deploying the most appropriate resource to enable timely response |
| 6. Resource location tracking | Requirements for determining the location of responding resources |
| 7. Patient data capture & destination triage | Hardware, software, and other essential products and technologies to record patient information at the scene and determine suitable hospital/facility |
| 8. Ad hoc communication | Requirements for real time communication between dispatch, deployed resources, callers, facilities, and others necessary to coordinate response and provide updates |
| 9. Information & research | Hardware, software, other essential products and technologies, and know-how needed to collect and use data for service improvement |
| 10. Human resources & training | Requirements for securing and maintaining the workforce for call-taking, resource dispatch, and communications in EMDS, including appropriate staffing models, training, supervision, and ongoing competency development |
| 11. Governance & leadership | Requirements for ensuring oversight, accountability, and strategic direction |

### 2. Delphi Survey

We identified and approached 68 potential participants; of these, 43 participated in the first round, 33 in the second round, and 30 in the third. Characteristics of participants are shown in Table 2.

**Table 2.**
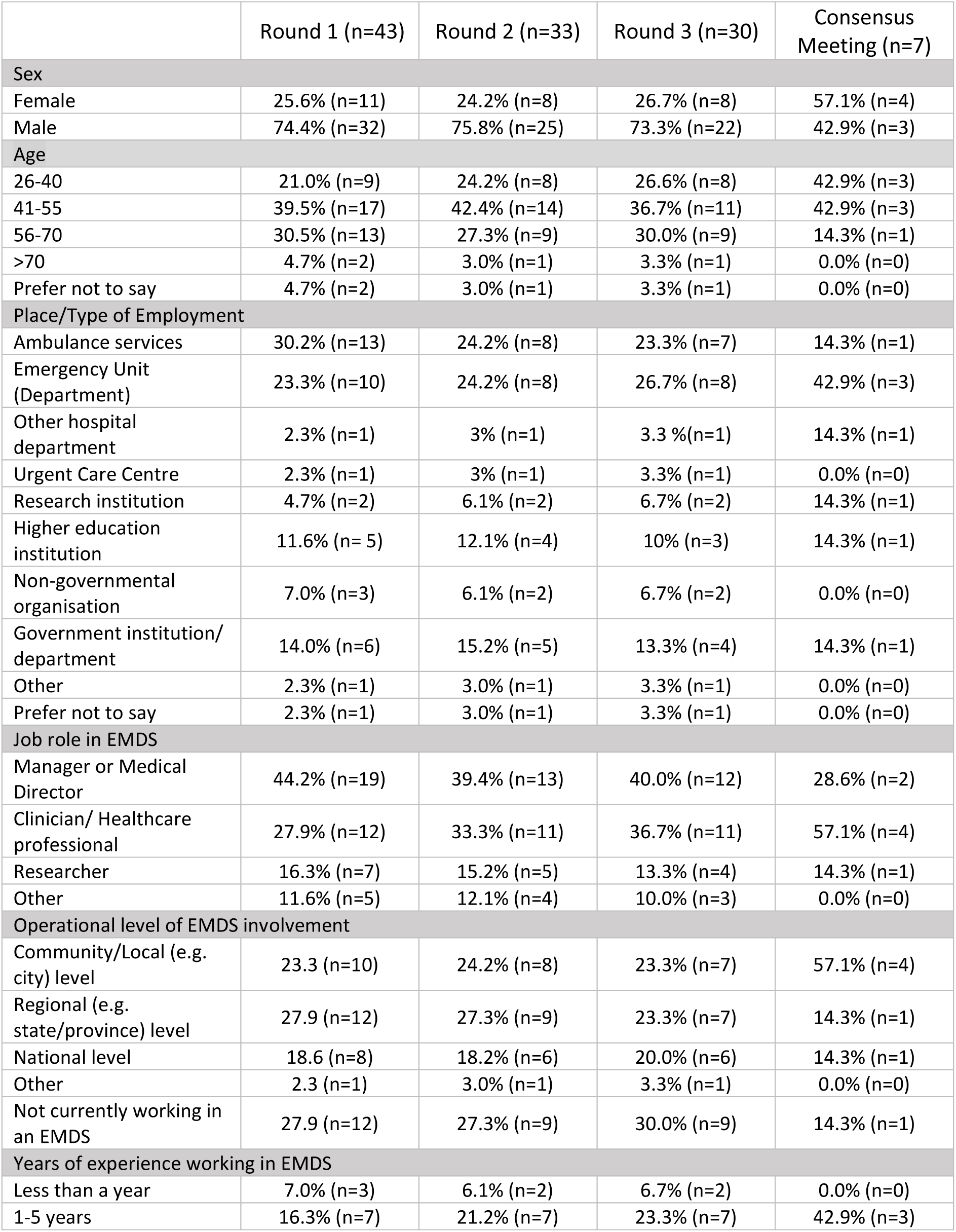
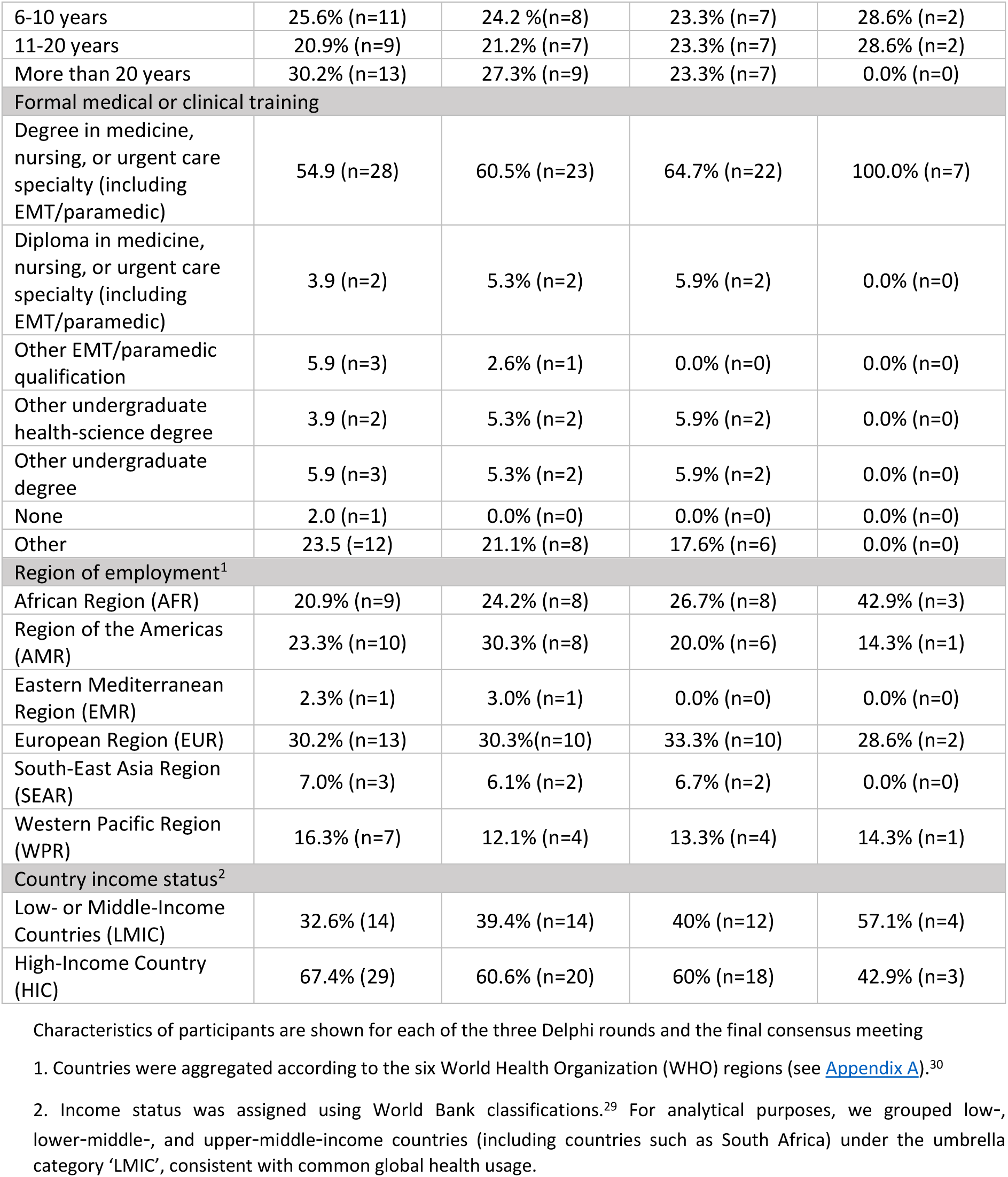
Participant Characteristics.

|  | Round 1 (n=43) | Round 2 (n=33) | Round 3 (n=30) | Consensus Meeting (n=7) |
| --- | --- | --- | --- | --- |
| **Sex** |  |  |  |  |
| Female | 25.6% (n=11) | 24.2% (n=8) | 26.7% (n=8) | 57.1% (n=4) |
| Male | 74.4% (n=32) | 75.8% (n=25) | 73.3% (n=22) | 42.9% (n=3) |
| **Age** |  |  |  |  |
| 26-40 | 21.0% (n=9) | 24.2% (n=8) | 26.6% (n=8) | 42.9% (n=3) |
| 41-55 | 39.5% (n=17) | 42.4% (n=14) | 36.7% (n=11) | 42.9% (n=3) |
| 56-70 | 30.5% (n=13) | 27.3% (n=9) | 30.0% (n=9) | 14.3% (n=1) |
| >70 | 4.7% (n=2) | 3.0% (n=1) | 3.3% (n=1) | 0.0% (n=0) |
| Prefer not to say | 4.7% (n=2) | 3.0% (n=1) | 3.3% (n=1) | 0.0% (n=0) |
| **Place/Type of Employment** |  |  |  |  |
| Ambulance services | 30.2% (n=13) | 24.2% (n=8) | 23.3% (n=7) | 14.3% (n=1) |
| Emergency Unit (Department) | 23.3% (n=10) | 24.2% (n=8) | 26.7% (n=8) | 42.9% (n=3) |
| Other hospital department | 2.3% (n=1) | 3% (n=1) | 3.3% (n=1) | 14.3% (n=1) |
| Urgent Care Centre | 2.3% (n=1) | 3% (n=1) | 3.3% (n=1) | 0.0% (n=0) |
| Research institution | 4.7% (n=2) | 6.1% (n=2) | 6.7% (n=2) | 14.3% (n=1) |
| Higher education institution | 11.6% (n= 5) | 12.1% (n=4) | 10% (n=3) | 14.3% (n=1) |
| Non-governmental organisation | 7.0% (n=3) | 6.1% (n=2) | 6.7% (n=2) | 0.0% (n=0) |
| Government institution/ department | 14.0% (n=6) | 15.2% (n=5) | 13.3% (n=4) | 14.3% (n=1) |
| Other | 2.3% (n=1) | 3.0% (n=1) | 3.3% (n=1) | 0.0% (n=0) |
| Prefer not to say | 2.3% (n=1) | 3.0% (n=1) | 3.3% (n=1) | 0.0% (n=0) |
| **Job role in EMDS** |  |  |  |  |
| Manager or Medical Director | 44.2% (n=19) | 39.4% (n=13) | 40.0% (n=12) | 28.6% (n=2) |
| Clinician/ Healthcare professional | 27.9% (n=12) | 33.3% (n=11) | 36.7% (n=11) | 57.1% (n=4) |
| Researcher | 16.3% (n=7) | 15.2% (n=5) | 13.3% (n=4) | 14.3% (n=1) |
| Other | 11.6% (n=5) | 12.1% (n=4) | 10.0% (n=3) | 0.0% (n=0) |
| **Operational level of EMDS involvement** |  |  |  |  |
| Community/Local (e.g. city) level | 23.3 (n=10) | 24.2% (n=8) | 23.3% (n=7) | 57.1% (n=4) |
| Regional (e.g. state/province) level | 27.9 (n=12) | 27.3% (n=9) | 23.3% (n=7) | 14.3% (n=1) |
| National level | 18.6 (n=8) | 18.2% (n=6) | 20.0% (n=6) | 14.3% (n=1) |
| Other | 2.3 (n=1) | 3.0% (n=1) | 3.3% (n=1) | 0.0% (n=0) |
| Not currently working in an EMDS | 27.9 (n=12) | 27.3% (n=9) | 30.0% (n=9) | 14.3% (n=1) |
| **Years of experience working in EMDS** |  |  |  |  |
| Less than a year | 7.0% (n=3) | 6.1% (n=2) | 6.7% (n=2) | 0.0% (n=0) |
| 1-5 years | 16.3% (n=7) | 21.2% (n=7) | 23.3% (n=7) | 42.9% (n=3) |
| 6-10 years | 25.6% (n=11) | 24.2% (n=8) | 23.3% (n=7) | 28.6% (n=2) |
| 11-20 years | 20.9% (n=9) | 21.2% (n=7) | 23.3% (n=7) | 28.6% (n=2) |
| More than 20 years | 30.2% (n=13) | 27.3% (n=9) | 23.3% (n=7) | 0.0% (n=0) |
| **Formal medical or clinical training** |  |  |  |  |
| Degree in medicine, nursing, or urgent care specialty (including EMT/paramedic) | 54.9 (n=28) | 60.5% (n=23) | 64.7% (n=22) | 100.0% (n=7) |
| Diploma in medicine, nursing, or urgent care specialty (including EMT/paramedic) | 3.9 (n=2) | 5.3% (n=2) | 5.9% (n=2) | 0.0% (n=0) |
| Other EMT/paramedic qualification | 5.9 (n=3) | 2.6% (n=1) | 0.0% (n=0) | 0.0% (n=0) |
| Other undergraduate health-science degree | 3.9 (n=2) | 5.3% (n=2) | 5.9% (n=2) | 0.0% (n=0) |
| Other undergraduate degree | 5.9 (n=3) | 5.3% (n=2) | 5.9% (n=2) | 0.0% (n=0) |
| None | 2.0 (n=1) | 0.0% (n=0) | 0.0% (n=0) | 0.0% (n=0) |
| Other | 23.5 (=12) | 21.1% (n=8) | 17.6% (n=6) | 0.0% (n=0) |
| **Region of employment<sup>1</sup>** |  |  |  |  |
| African Region (AFR) | 20.9% (n=9) | 24.2% (n=8) | 26.7% (n=8) | 42.9% (n=3) |
| Region of the Americas (AMR) | 23.3% (n=10) | 30.3% (n=8) | 20.0% (n=6) | 14.3% (n=1) |
| Eastern Mediterranean Region (EMR) | 2.3% (n=1) | 3.0% (n=1) | 0.0% (n=0) | 0.0% (n=0) |
| European Region (EUR) | 30.2% (n=13) | 30.3% (n=10) | 33.3% (n=10) | 28.6% (n=2) |
| South-East Asia Region (SEAR) | 7.0% (n=3) | 6.1% (n=2) | 6.7% (n=2) | 0.0% (n=0) |
| Western Pacific Region (WPR) | 16.3% (n=7) | 12.1% (n=4) | 13.3% (n=4) | 14.3% (n=1) |
| **Country income status<sup>2</sup>** |  |  |  |  |
| Low- or Middle-Income Countries (LMIC) | 32.6% (14) | 39.4% (n=14) | 40% (n=12) | 57.1% (n=4) |
| High-Income Country (HIC) | 67.4% (29) | 60.6% (n=20) | 60% (n=18) | 42.9% (n=3) |

Characteristics of participants are shown for each of the three Delphi rounds and the final consensus meeting
1. Countries were aggregated according to the six World Health Organization (WHO) regions (see Appendix A).<sup>30</sup> 2. Income status was assigned using World Bank classifications.<sup>29</sup> For analytical purposes, we grouped low-, lower-middle-, and upper-middle-income countries (including countries such as South Africa) under the umbrella category 'LMIC', consistent with common global health usage.

#### Delphi outcomes

After Round 1, 36 additional components were proposed for specific levels of EMDS, resulting in a total number of 369 components considered in the Delphi (333 from the initial expert workshop [111 at each level] and 36 additional at specific levels). In the main analysis, by Round 3, 289 (78.3%) components met the ≥75% threshold for inclusion in one or more EMDS levels, 18 (4.9%) met the ≥75% threshold for exclusion from all levels, and 62 did not reach consensus (Figure 1). Among the 289 components which reached agreement for inclusion at any EMDS level, 87 (23.6%) were in Foundational, 101 (27.4%) in Emerging, and 101 (27.4%) in established EMDS levels.

**Figure 1.**
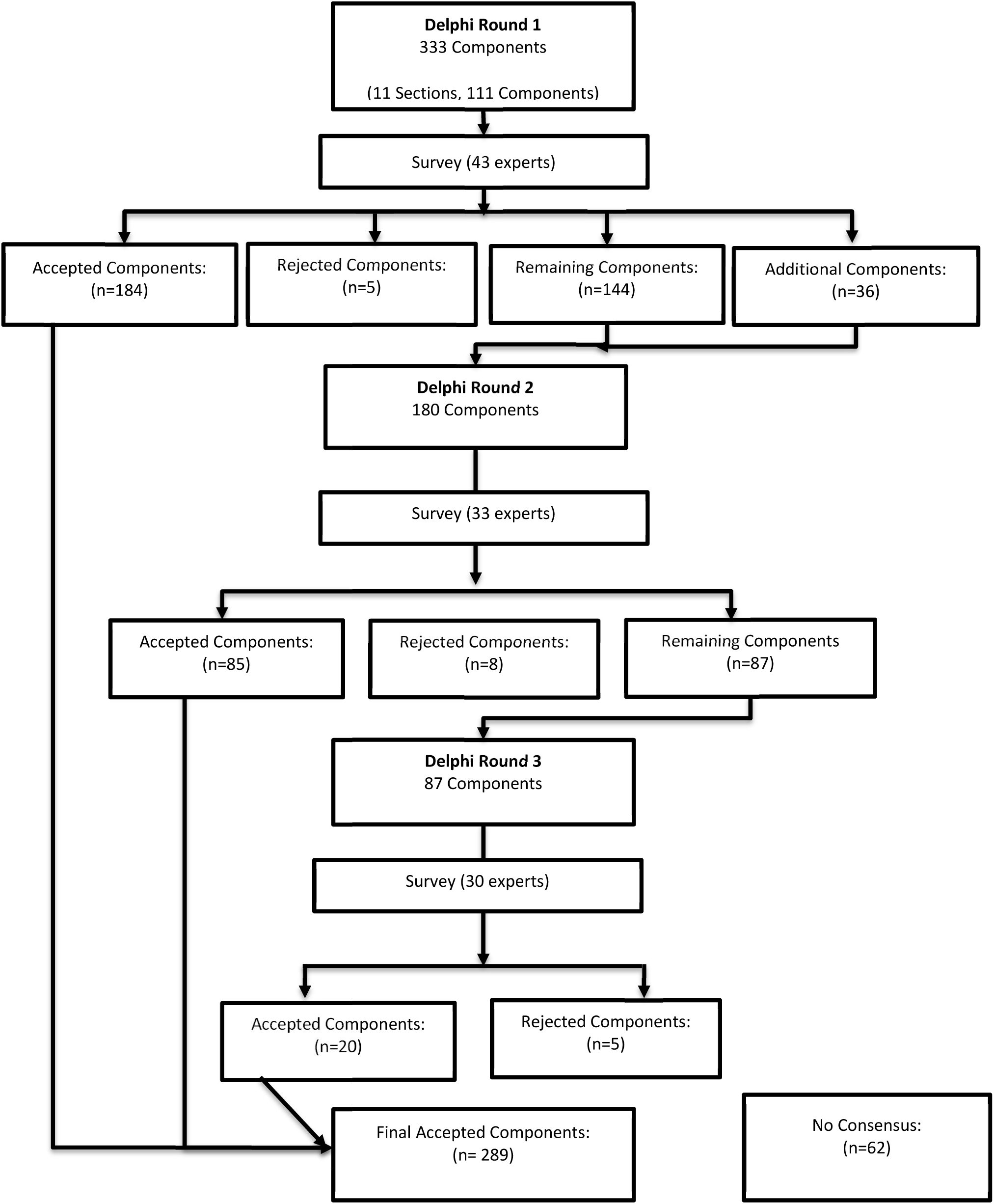
Delphi Flowchart Main Analysis.

Figure 1 shows the flow of the Delphi for the main analysis involving all participants (regardless of country income status). Results by country income status are presented in Appendices B and C.

#### Subgroup outcomes by income setting

For subgroup analyses restricted to participants whose primary country of work was a LMIC, after all three rounds, 318 (86.2%) components had agreement to be included across all levels (Appendix B). Of those 318 components, 105 (33.2%) were included at the Foundational level, 107 (33.6%) in Emerging, and 106 (33.3%) in Established. For analyses restricted to participants whose primary workplace was in a HIC, 257 (69.6%) components reached consensus for inclusion (see Appendix C). Of those 257 components, 72 (28.0 %) were included only in Foundational, 91 (35.4%) only in Emerging, and 94 (36.6%) only in Established.

There were 66 components which LMIC respondents agreed to include which HIC respondents did not; 34 of these were at the Foundational level and they tended to be technologically focussed, for example, including real-time electronic data capture tools, displaying screens in dispatch to show location of resources, use of emergency apps on mobile phones. There were 5 components which HIC respondents included but LMIC respondents did not, 4 of these were in Emerging or Established systems (Appendix E).

When deriving a list of components which should be recommended for each level of system, based on country income strata of respondents, 300 components were included; 105 (35.0%) were included in the Foundational, 101 (33.7%) in the Emerging, and 94 (31.3%) in the Established levels (Appendix F).

Given component’s inclusion in subsequent rounds was determined by the analysis done for all participants, it was possible that some components which would have been included in a subsequent round by each income strata group were not represented in that round. There were 9 such components identified. These components are marked with an asterisk in Appendix D and listed in full in Appendix G.

#### Consensus meeting

The consensus meeting was attended by 7 experts (4 from LMICs and 3 from HICs). Prior to the meeting, study leads (JD, LW, AI, AN, WS, SJ, and VN) appraised the results and identified responses for components which were inconsistent within and across levels – these included components which were incompatible (for example, real time electronic analysis of data, but use of paper forms to collect data) and those which seemed inconsistent across levels (for example, included in Foundational and Established but not Emerging). To resolve inconsistencies, discussions focused on the practicality and feasibility of proposed components within each domain and level. Several components that had been endorsed at the Foundational level in earlier analyses were particularly technologically advanced or infrastructure-dependent tools. These were re-evaluated, re-assigned to different levels, consolidated, or removed where feasibility constraints were thought to make them unsuitable as essential components for early-stage systems: e.g., advanced GPS, automated digital interfaces, resource-intensive communication tools. This process removed 42 components from Foundational, 17 from Emerging, and 15 from Established. It added one component to Established and resulted in a final list of 227 components across 11 domains across the three levels, including 63 at the Foundational, 84 at the Emerging, and 80 at the Established levels (Table 3).

**TABLE 3:** FINAL RESULTS.

| Domain | Component | Level |  |  |
| --- | --- | --- | --- | --- |
|  |  | Foundational | Emerging | Established |
| **Section 1: Essential products and technologies: Hardware and “high level” software, general system requirements – essential products and technologies** | 1.1 Phone-landline based communication system<sup>1</sup> | ● | ● | ● |
|  | 1.2 Radio communication system<sup>1</sup> | ● | ● | ● |
|  | 1.3 Mobile phone communication system<sup>1</sup> | ● | ● | ● |
|  | 1.4 Mobile phones or tablets in ambulances<sup>1</sup> | ● | ● | ● |
|  | 1.5 Simple paper-based data collection forms for use by dispatch staff (call takers and resource dispatchers) | ● |  |  |
|  | 1.6 Simple paper-based data collection forms for use by ambulance providers | ● |  |  |
|  | 1.7 Computer hardware | ● | ● | ● |
|  | 1.8 Standardised computer-aided dispatch software |  | ● | ● |
|  | 1.9 A central directory of caller numbers | ● | ● | ● |
|  | 1.10 Technologies to automatically identify and/or capture caller number and or address |  | ● | ● |
|  | 1.11 Ability to communicate manually (e.g. by radio or telephone) with other emergency agencies<sup>1</sup> | ● | ● | ● |
|  | 1.12 An electronic system for communication with other emergency services<sup>1</sup> |  | ● | ● |
|  | 1.13 An electronic system with individual location identification of other (non-ambulance) emergency services |  | ● | ● |
|  | 1.14 Screens at dispatch to show status of resources and accepted patients |  | ● | ● |
|  | 1.15. Screens at facility to show status of incoming patients |  | ● | ● |
|  | 1.16 Automated translation for non- native language speaking- callers (accessibility features and language translations) |  | ● | ● |
|  | 1.18 Non-voice based options for accessing the dispatch centre (e.g. SMS) for callers who are in danger when speaking, or people with hearing impairments |  |  | ● |
| **Section 2: Call Taking: Includes identifying the emergency and caller location** | 2.1. Standard emergency medical system call number(s) (e.g. 999 or 911) for respective geographic areas | ● | ● | ● |
|  | 2.3 An electronic data capture system for entering caller details, location, and details of the emergency at the end of the call | ● |  |  |
|  | 2.4 An electronic data capture system for entering caller details, location, and details of the emergency in real time |  | ● | ● |
|  | 2.5 A paper map-based method for identification of emergency location | • |  |  |
|  | 2.6 Bespoke (customised) mapping of local landmarks to guide in emergency location | • | • | • |
|  | 2.7 Software to enable sharing of the patient's "pin" location |  | • | • |
|  | 2.9 Electronic mapping software to aid in emergency location |  | • | • |
|  | 2.10 Cell phone triangulation to aid in emergency location |  | • | • |
|  | 2. 11 An emergency app on mobile phones (of the population of potential callers) with preloaded caller GPS to aid in emergency location |  | • | • |
|  | 2.8 "What three words" (or similar location tools) to aid in emergency location |  | • | • |
| **Section 3: Emergency Prioritisation (Triage): Assess urgency of call** | 3.1 An all or nothing system (any vs no ambulance response to all calls) | • |  |  |
|  | 3.4 Criteria-based prioritisation - based on a clinically qualified call taker perception of urgency<sup>1</sup> |  | • |  |
|  | 3.6 Computer-based medical priority dispatch software (questions and computer-algorithms to prioritise which patients should be attended by an ambulance)<sup>1</sup> |  | • | • |
| **Section 4: Advice Giving: Provide immediate guidance to the caller** | 4.1 Patient complaint-based, non-scripted, simple instructions<sup>1</sup> | • | • |  |
|  | 4.2 Patient complaint-based, non-scripted, intervention coaching<sup>1</sup> | • | • | • |
|  | 4.3 Patient complaint-based, scripted, simple instructions<sup>1</sup> | • | • | • |
|  | 4.4 Patient complaint-based, scripted, intervention coaching<sup>1</sup> | • | • | • |
|  | 4.5 Photograph-assisted advice | • | • |  |
|  | 4.7 Linking the patient to primary care for advice | • | • | • |
| **Section 5: Ambulance Dispatch: Assign appropriate resource (ambulance or first responder)** | 5.1 Manual (paper or electronic) reporting and recording of resource location in relation to incident location | • |  |  |
|  | 5.3 Regular manual (paper or electronic) recording of resource status (e.g.: on readiness to attend an incident) | • |  |  |
|  | 5.4 Automatic (electronic) system to update on resource status (e.g.: on readiness to attend an incident) |  | • | • |
|  | 5.8 Allocation of the closest available resource to attend emergencies, regardless of patient acuity or incident type | • |  |  |
|  | 5.5. A policy of "resource return to basecamp"<sup>2</sup> | • | • |  |
|  | 5.6 Policy of "resource return to pre-identified standby location" | • | • | • |
|  | 5.2 GPS-enabled electronic-tracking system of ambulances |  | • | • |
|  | 5.7 Dynamic resource relocation of ambulances and crews<sup>3</sup> |  | • | • |
|  | 5.9 Prioritisation of resource type (level of equipment /staffing of ambulance) in accordance with patient acuity or incident type |  | • | • |
|  | 5.11 Prioritisation of resource based on a clinician's assessment of patient's need |  | • |  |
|  | 5.13 Prioritisation of resource type should be computer-based (e.g.: using algorithms) |  | • | • |
| **Section 6: Emergency Location Tracking & ambulance routing: Monitor ambulance *en route* to the emergency** | 6.1 Ambulance crew location tracking based on verbal description of emergency location | • |  |  |
|  | 2.12 GPS location mapping and routing function for responding resources |  | • | • |
|  | 2.13 GPS location mapping via vehicle-based GPS system<sup>1</sup> |  | • | • |
|  | 2.14 GPS location mapping via phone-based app that can provide GPS<sup>1</sup> |  | • | • |
| **Section 7: Patient Data Capture & Destination Triage: Record patient info and determine suitable hospital** | 7.1 At the scene, information on the patient and their condition captured on bespoke paper forms | • |  |  |
|  | 7.2 At the scene, information on the patient and their condition captured into an electronic interface |  | • | • |
|  | 7.4 Destination triage based on the predesignated facilities' ability to treat the patient's condition, considering the patient's acuity or the type of emergency | • | • | • |
|  | 7.5 Destination triage to predesignated facilities, based on their ability to treat the patient's condition, based on use of paper-based flow charts | • |  |  |
|  | 7.6 Destination triage to predesignated facilities, based on their ability to treat the patient's condition, is computer aided (based on patient data and algorithms) |  | • | • |
|  | 7.7 Destination triage altered dynamically, based on the readiness of predesignated facilities to receive patients <sup>5</sup> | • | • | • |
| **Section 8: Dynamic communication between dispatch, crew, and callers** | 8.2 Communication between any or all of dispatch, crews or facilities to be verbal and scripted (or written down/protocolised)<sup>1</sup> | • | • | • |
|  | 8.3 Electronic patient data captured at dispatch to be sent to devices in ambulances |  | • | • |
|  | 8.4 Electronic patient data captured by ambulances to be sent to devices in facilities and/or dispatch |  | • | • |
| **Section 9: Information & Research: Support decision-making and continuous improvement** | 9.1 Collection and archive of basic standardised paper data capture sheets in dispatch and ambulances (including any or all of caller and emergency details, advice given to caller, resource prioritisation, resource status and use, emergency details [from the scene], and destination triage) | • |  |  |
|  | 9.3 Paper data regularly input into an electronic file | • |  |  |
|  | 9.4 Real-time (electronic) data capture (including any or all of caller and emergency details, resource prioritisation, advice given to caller, resource status and use, emergency details [from the scene], and destination triage) |  | • | • |
|  | 9.12 Recording capability of phone and radio conversations, resource deployment tools, and evidence-based call evaluation software |  |  | • |
|  | 9.6 Regular retrospective data analysis (including any or all of caller and emergency details, resource prioritisation, advice given to caller, resource status and use, emergency details [from the scene], and destination triage) | • | • | • |
|  | 9.8 On demand/ad hoc data-informed quality improvement meetings (including on procedures at dispatch, in ambulances, and of destination triage) | • | • | • |
|  | 9.9. Regular data-informed quality improvement meetings (including on procedures at dispatch, in ambulances, and of destination triage) | • | • | • |
| **Section 10: Human Resources & Training: Ensure skilled staff and ongoing development** | 10.1 Dispatch staff both receive calls and dispatch resources<sup>4</sup> | • | • | • |
|  | 10.6 Staff receiving calls in dispatch have standardised basic training (based on tools or systems being used in call taking and communication) | • | • | • |
|  | 10.7 Staff dispatching resources have standardised basic training (to the level of tools or systems which are deployed in resource dispatch and communication) | • | • | • |
|  | 10.19 Mandatory credentialling (certification at national level) for dispatch staff (call takers and or resource dispatchers) appropriate to level of EMDS |  | • | • |
|  | 10.4 Dedicated ambulance providers who are tasked with assessing and providing care to patients | • | • | • |
|  | 10.8 Ambulance providers have standardised basic training (in patient assessment and care provision) | • | • | • |
|  | 10.16 Ambulance providers have a bespoke qualification (degree, diploma or equivalent) |  | • | • |
|  | 10.13 Ambulance providers are clinically trained (nurse, doctor, or paramedic/EMT) | • | • | • |
|  | 10.17 Mandatory credentialling (certification at national level) for ambulance providers appropriate to level of EMDS |  | • | • |
|  | 10.18 Any or all of call takers, resource dispatchers, or ambulance crews (EMT/paramedics) are trained in basic data analysis and reporting |  | • | • |
|  | 10.5 Dedicated staff in facilities to receive calls | • | • | • |
|  | 10.9 Staff receiving calls in facilities have standardised basic training (to the level of tools which are deployed in decisions to accept patients at facilities) | • | • | • |
|  | 10.12 Staff receiving calls in facilities are clinically trained (nurse, doctor, or paramedic/EMT) | • | • | • |
| **Section 11: Governance & Leadership: Oversight, accountability, and strategic direction** | 11.1 Off-site operational oversight (e.g.: a chief operating officer [COO]) | • | • |  |
|  | 11.2 On-site operational oversight (e.g.: a chief operating officer [COO]) | • | • | • |
|  | 11.3 Operational oversight of dispatch staff is distinct (a separate person/role) to that of ambulance providers | • | • | • |
|  | 11.4 Clinically trained staff providing operational oversight of dispatch staff (call takers or resource dispatchers) | ● | ● | ● |
|  | 11.5 Clinically trained staff providing operational oversight of ambulance providers | ● | ● | ● |
|  | 11.18 Regular meetings between emergency medical services and facility leadership to discuss referral pathways and destination triage |  | ● | ● |
|  | 11.6 SOPs for processes involved in all of call taking, emergency location, and resource triage | ● | ● | ● |
|  | 11.7 SOPs for processes involved in resource location and status maintenance | ● | ● | ● |
|  | 11.8 SOPs on destination triage | ● | ● | ● |
|  | 11.9 SOPs for minimum data capture by call takers and resource dispatchers | ● | ● | ● |
|  | 11.10 SOPs for minimum data capture by ambulance providers | ● | ● | ● |
|  | 11.11 SOPs for communicating between EMS personnel | ● | ● | ● |
|  | 11.12 SOPs for giving advice to callers/patients | ● | ● | ● |
|  | 11.13 SOPs for data analysis | ● | ● | ● |
|  | 11.14 SOPs for data reporting | ● | ● | ● |
|  | 8.5 Detailed protocols (including for disaster/mass casualty) | ● | ● | ● |
|  | 11.15 Data security protocols (data protection and privacy), which are enforced | ● | ● | ● |
|  | 11.16 Transparent reporting of analysed data to public and relevant stakeholders | ● | ● | ● |
|  | 11.17 Clear policies on risk appetite and acceptable over- and under-triage rates | ● | ● | ● |
| **Total** |  | **63** | **84** | **80** |

<sup>1</sup> The presence of multiple similar options presented as basic requirements reflects ambivalence during the consensus meeting about what should constitute the minimum requirement.
<sup>2</sup> The recommendation is that, although a return to basecamp is endorsed as the basic requirement for the Foundational and Emerging levels, in special events this should be replaced by provision for ambulances to go directly to specific locations.
<sup>3</sup> Although dynamic resource relocation is not supported at the Foundational level, planned relocation for special events does occur in practice and may be appropriate where capacity and coordination permit.
<sup>4</sup> Although this is a minimum requirement for the Established level, it is recommended that the roles be fulfilled by separate individuals, depending on the context.
<sup>5</sup> Noting that, for Foundational systems, this refers to dynamic alteration of destination decisions when there are known issues with, for example, availability of essential equipment's. This doesn't refer to continuous destination triage, which requires computerised data collection at facilities.

## DISCUSSION

This modified Delphi study established consensus on essential components required for EMDS across diverse levels of development. Through iterative expert consultation, participants endorsed a comprehensive framework spanning governance, communication infrastructure, operational processes, and technological capabilities. Final agreement reflected a consistent levelled pattern across domains, with Foundational components primarily focused on governance and basic communication functions, and more complex technologies and protocols more frequently positioned within the Emerging and Established levels. The resulting framework offers a globally relevant structure that accommodates diverse resource contexts while maintaining alignment with core principles of emergency medical dispatch.

The Delphi process generated strong consensus on essential EMDS components, with many items reaching agreement in rounds one and two. Agreement was consistently high for components related to communication systems, mapping and navigation, structured call-taking and intervention coaching, resource tracking, and data management. LMIC respondents endorsed more components overall, and more at the Foundational level, than HIC respondents, likely reflecting differences in context and operational priorities rather than divergent views on importance.

Items failing to reach consensus related to technologically or organisationally complex components often with more LMIC than HIC participants stating these should be components of Foundational systems. As a result of the final expert consensus meeting, clearer differentiation emerged across the three maturity levels, with the Foundational level streamlined to reflect a minimal, feasible set of core functions rather than a comprehensive or highly detailed system. The resulting components should be feasible for less well-resourced or technologically advanced settings and are pragmatically aligned with system maturity expectations, whilst preserving the core functions with the strongest participant endorsement.

The pattern of endorsement underscored the centrality of reliable communication, structured call-taking, operational clarity, and pragmatic data capture as the bedrock of EMDS. However, the placement of several technologically advanced components at the Foundational level, predominately by LMIC respondents, may highlight a gap between desirability and feasibility in system development. This may also reflect both perceived efficiency gains and the rapidly evolving technological landscape across many low-resource settings - where digital solutions could be used to leapfrog persistent analogue constraints, or weak or inconsistent infrastructure (e.g., fragmented radio networks, variable paper data flows).^31^ ^32^ With regards to LMICs, given the rapid expansion of mobile connectivity and digital technology adoption in many countries, progression toward technology-enabled system components may be rapid once infrastructure and affordability constraints are addressed.^31^ ^32^ However, existing literature provides limited evidence on the successful scale-up and long-term sustainability of highly complex, fully integrated dispatch systems in most LMIC settings, with published analyses instead emphasising persistent constraints related to infrastructure, workforce capacity, financing, and system integration.^1^ ^15^ ^23^ In contexts with limited resources, early adoption of advanced technologies may therefore introduce challenges related to alignment with local system realities and workforce capacity.

Nevertheless, the three levels should be understood as markers of system capability rather than proxies for national income classification. Some high-income contexts continue to operate relatively analogue dispatch systems ^33^ ^34^, particularly in remote or low-volume settings or where incentives are not there for efficiency or change,^34^ ^35^ whilst some low- and middle-income settings implement sophisticated digital solutions from the outset because they directly address critical operational barriers.^36^ ^37^ The consensus in this Delphi supports a staged approach starting with core functions, depending on needs and resource availability. Technology should then be introduced where it genuinely strengthens reliability, safety, and coordination (cf. WHO Emergency Care System Framework and broader health system guidance).^21^ This interpretation is consistent with the limited literature describing emergency dispatch and prehospital coordination in low-resource contexts in LMICs, where simplified triage processes and basic communication systems are often highlighted as pragmatic approaches for maintaining system function under resource constraints.^5^ ^16^ ^21–23^

## STRENGTHS AND LIMITATIONS

A key strength is the breadth of expertise and geographical representation captured through a structured, transparent process, complemented by an expert consensus meeting to reconcile overlaps and ensure coherent level placement. The framework provides a pragmatic roadmap for establishing and advancing EMDS, supporting countries to begin with essential governance and communication functions and to layer technology and protocols as capability grows. It aligns with global policy priorities, including the World Health Assembly resolution on Integrated Emergency, Critical and Operative Care (WHA76.2), which calls for coherent, context appropriate guidance to reduce preventable delays in care.^20^ ^21^ The levelled structure also lends itself to benchmarking and staged implementation planning, including the development of indicators for system performance, training, and quality improvement, while recognising that progression across levels is not strictly linear and that different system elements may develop at different rates.

Several limitations warrant consideration. First, subgroup-specific scoring during the Delphi survey was not feasible at the outset; consequently, some items progressed or were removed based on the overall participant responses before LMIC/HIC perspectives could be analysed separately. We addressed this through a retrospective analysis to identify items that were removed early. These components have been clearly highlighted in our analysis. Second, the imbalance in respondent numbers (HIC n=29 vs LMIC n=14) may have weighted the overall (all-participant) consensus process towards HIC perspectives in earlier rounds before subgroup analyses were applied. Third, although feasibility considerations may have been implicitly incorporated by some participants, this was not assessed directly, and Delphi responses may reflect expert judgement alongside aspirational components that do not fully account for feasibility, cost, sustainability, or workforce constraints. Fourth, interpretation of the level labels (Foundational, Emerging, Established) may have varied between respondents despite provision of level descriptors, and some participants, particularly those with limited exposure to EMDS models outside their own setting, may have interpreted “levelling” as reflecting system maturity, resource availability, or technological sophistication in different ways. Finally, consensus thresholds are useful for identifying agreement but do not capture implementation complexity or interdependencies. For instance, high-tech items may require stable connectivity, reliable electricity, substantial financial investment, and maintenance of supply chains not widely available in all settings. These caveats suggest cautious interpretation of endorsements that are technology dependent at Foundational level, and the need for context-specific staging. To mitigate this, level definitions were grounded in incremental capability (core governance/communication first, then progressively more complex protocols and technologies), and the post-Delphi consensus meeting explicitly reviewed level placement to ensure internal coherence and practical staging across settings.

## CONCLUSION

This study provides the first global consensus on essential components for Prehospital Emergency Medical Dispatch Systems, offering a levelled framework that can guide policy, planning, and benchmarking across diverse resource contexts and levels of emergency care system development. Future efforts should focus on operationalising this framework into practical tools – such as adaptable call taking scripts and coaching prompts, scalable data capture and destination triage templates, and implementation, and equity-focused indicators. Prospective pilot studies are essential to validate feasibility and refine level placement. Aligning these results with WHO frameworks and establishing periodic updates (for example, Utstein style convenings^38^ ^39^ for chain of survival related consensus) will ensure the guidance remains scalable, evidence-based, and responsive to evolving technology.

### COLLABORATORS: DISPATCH GUIDANCE WORKING GROUP

Naseef Abdullah, Hamidreza Aghababaeian, Motunrayo Ajisola, Alexei A. Birkun, Joe Bonney, Florian Breuer, Sharon Anoush Chekijian, Rafael Castro Delgado, Chip Decker, Agron Dogjani, Georgette Eaton, Faith Joan Gaerlan, James J Garland, Adish Gautam, Heike Geduld, Leila Ghalichi, Mo Ibrahim, Joseph K Kalanzi, Jo Kramer-Johansen, Justine Lee, Chih-Hao Lin, Koenraad G. Monsieurs, Felistus Ndanu Musyoka, Maxwell Osei-Ampofo, Jerry Overton, G.V. Ramana Rao, Karen Smith, Emnet Tesfaye Shimber, Jonathan D. Washko, David K. Whitaker, Erik Zakariassen

## Data Availability

All data produced in the present study are available upon reasonable request to the authors.

## ACKNOWLEDGMENTS

We sincerely thank all Delphi participants and expert panel members for their time, expertise, and thoughtful contributions, without which this work would not have been possible. This study was supported by funding from the Laerdal Foundation (Project number: Project, 2024-02950). The funders had no role in the study design, data collection, analysis, interpretation, or writing of the manuscript.

Supplementary Materials

## Appendix A: Participant Country list and income group

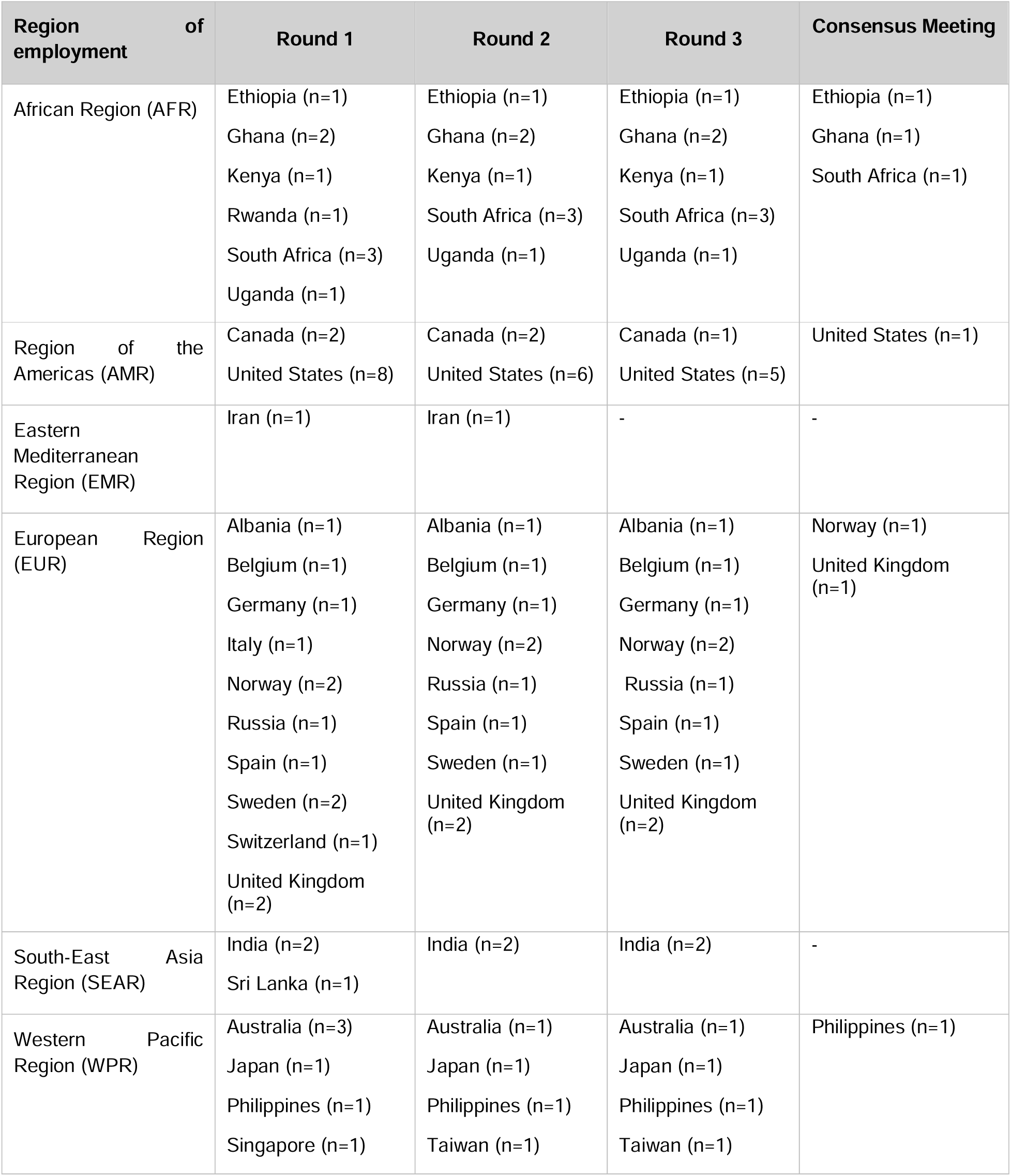
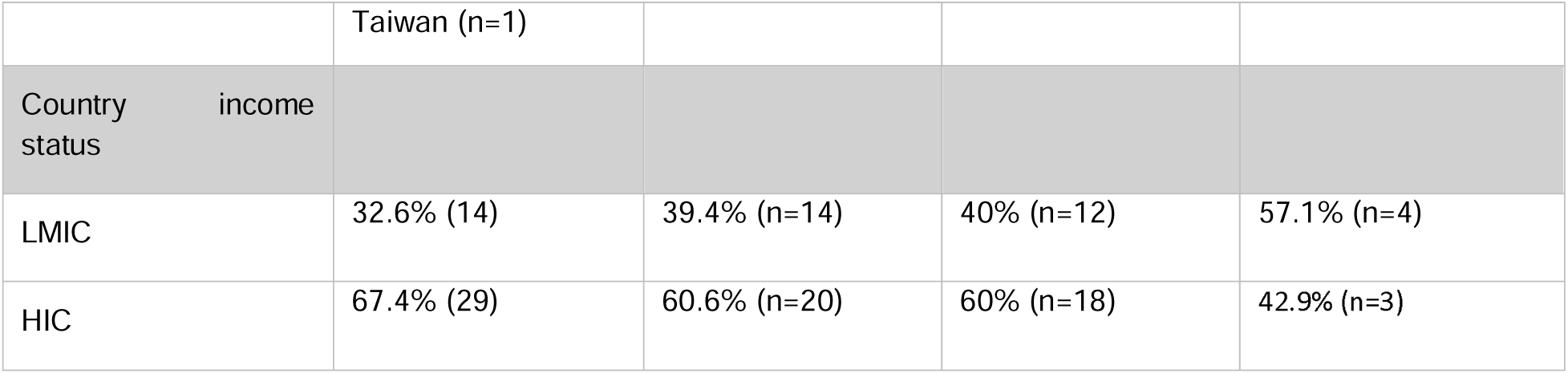

## Appendix B: Subgroup Analysis: LMIC Participants

For the analysis including only participants whose primary country of work was an LMIC, after round 1, 234 of the original 333 components were removed from subsequent rounds; 230 of these reached agreement to be included in all levels, 58 reached agreement to be included in Foundational, 88 in Emerging, and 84 in Established; 4 were removed from any level. In round 2, participants selected from 135 components, including the 36 which had been suggested in round 1 and the 99 components which remained from this round. After round 2, 74 components were removed; 68 of these reached agreement to be included in all levels, additionally 35 were agreed to be included only in Foundational, 15 only in Emerging, and 18 only in Established; 6 were removed from any level. In round 3, 31 components were removed; 20 reached agreement to be included in all levels, additionally 12 reached agreement to be included only in Foundational, 4 only in Emerging, and 4 only in Established; 11 were removed from any level. After all three rounds, of the 369 components proposed, 318 (86.2%) had agreement to be included. Of those 318 reaching agreement across all levels, 105 (33.2%) only in Foundational, 107 (33.6%) only in Emerging, and 106 (33.3%) only in Established (Appendix Figure 2 and Appendix F).

**Appendix Figure 2:**
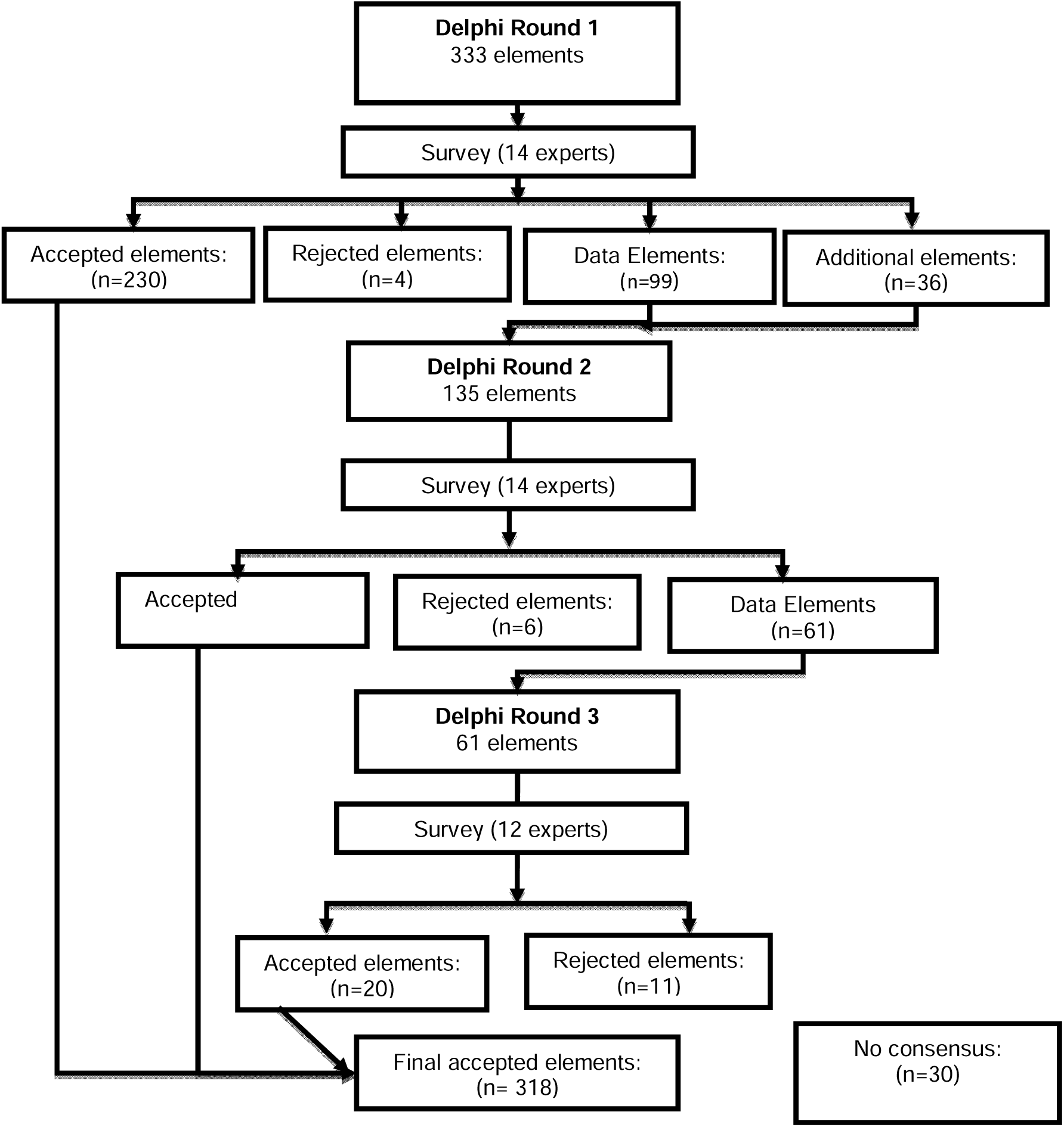
Delphi Flowchart for LMIC participants.

## Appendix C: Subgroup Analysis: HIC Participants

**Appendix Figure 3:**
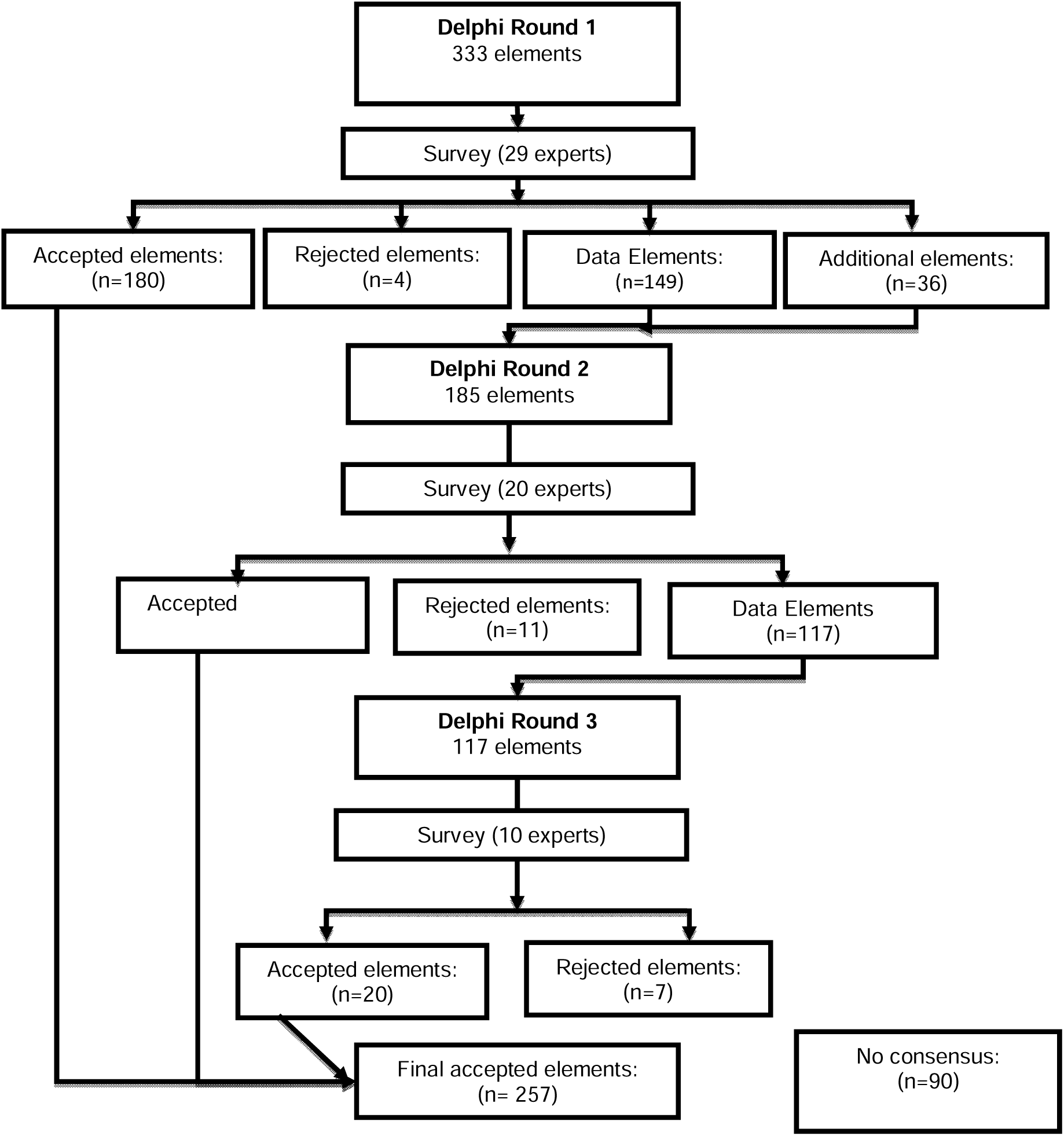
Delphi Flowchart HIC participants.

For the analysis including participants with their main workplace in a HIC, after round 1, 184 of the original 333 components were removed; 180 of these reached agreement to be included in all levels, with 35 reached agreement to be included only in Foundational, 71 only in Emerging, and 74 only in Established; 4 were removed from any level. In Round 2, participants were presented with 185 (including the 36 which had been suggested in round 1 and the 149 components which remained from this round. After round 2, 68 components were removed; 57 of these reached agreement to be included in all levels, 25 were agreed to be included only in Foundational, 16 only in Emerging, and 16 only in Established; 11 were removed from any level. In Round 3, 27 components were removed leaving 20 which reached agreement to be included in all levels, of which 12 reached agreement to be included only in Foundational, four only in Emerging, and four only in Established; 7 were removed from any level. After all three rounds, of the 369 components proposed (the initial 333 plus 36 additional suggestions), 257 (69.6%) components reached agreement to be included, 72 (28.0 %) were included only in Foundational, 91 (35.4%) only in Emerging, and 94 (36.6%) only in Established (Appendix Figure 3 and Appendix F).

## Appendix D: Final Delphi Results After Subgroup Analysis

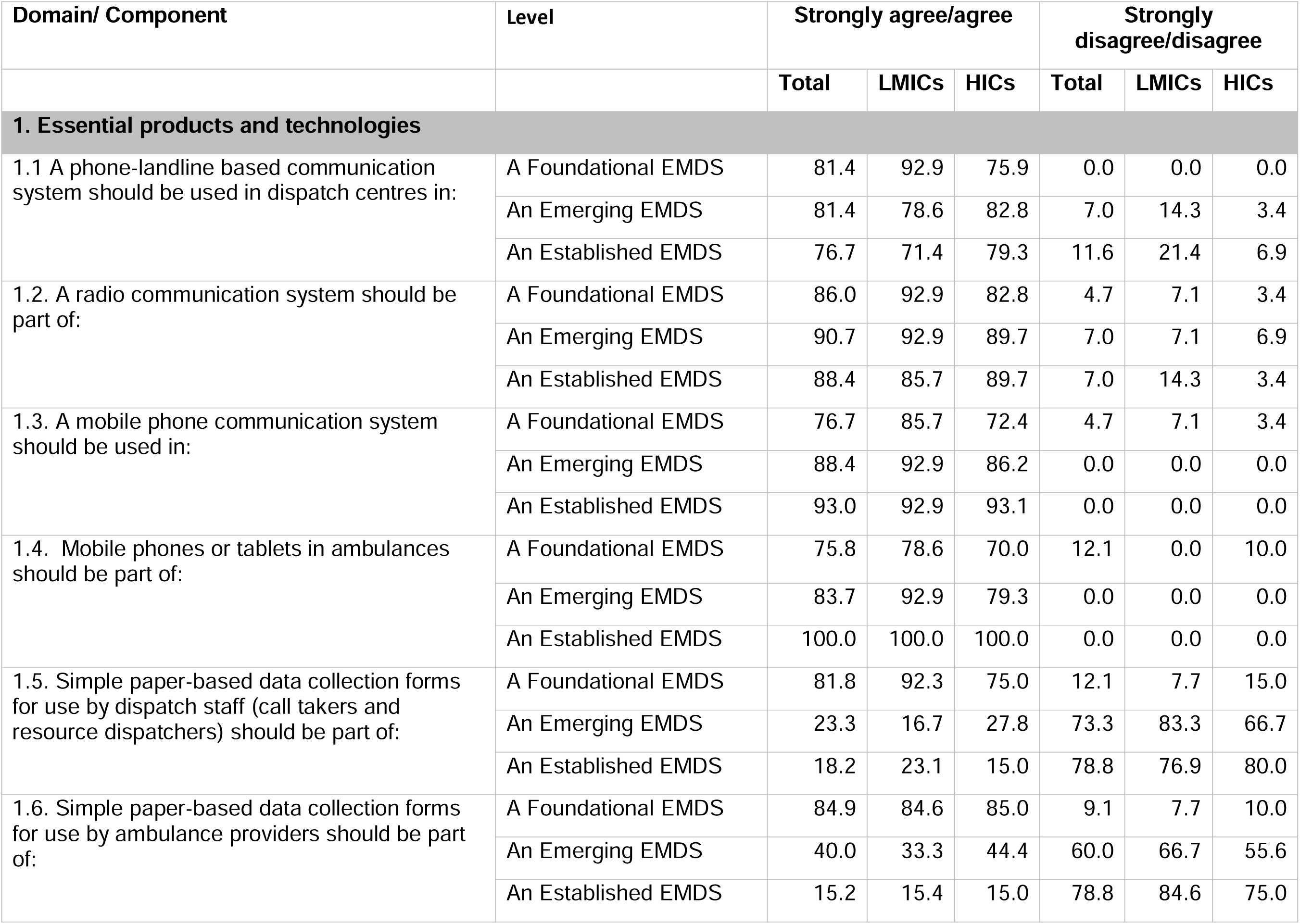
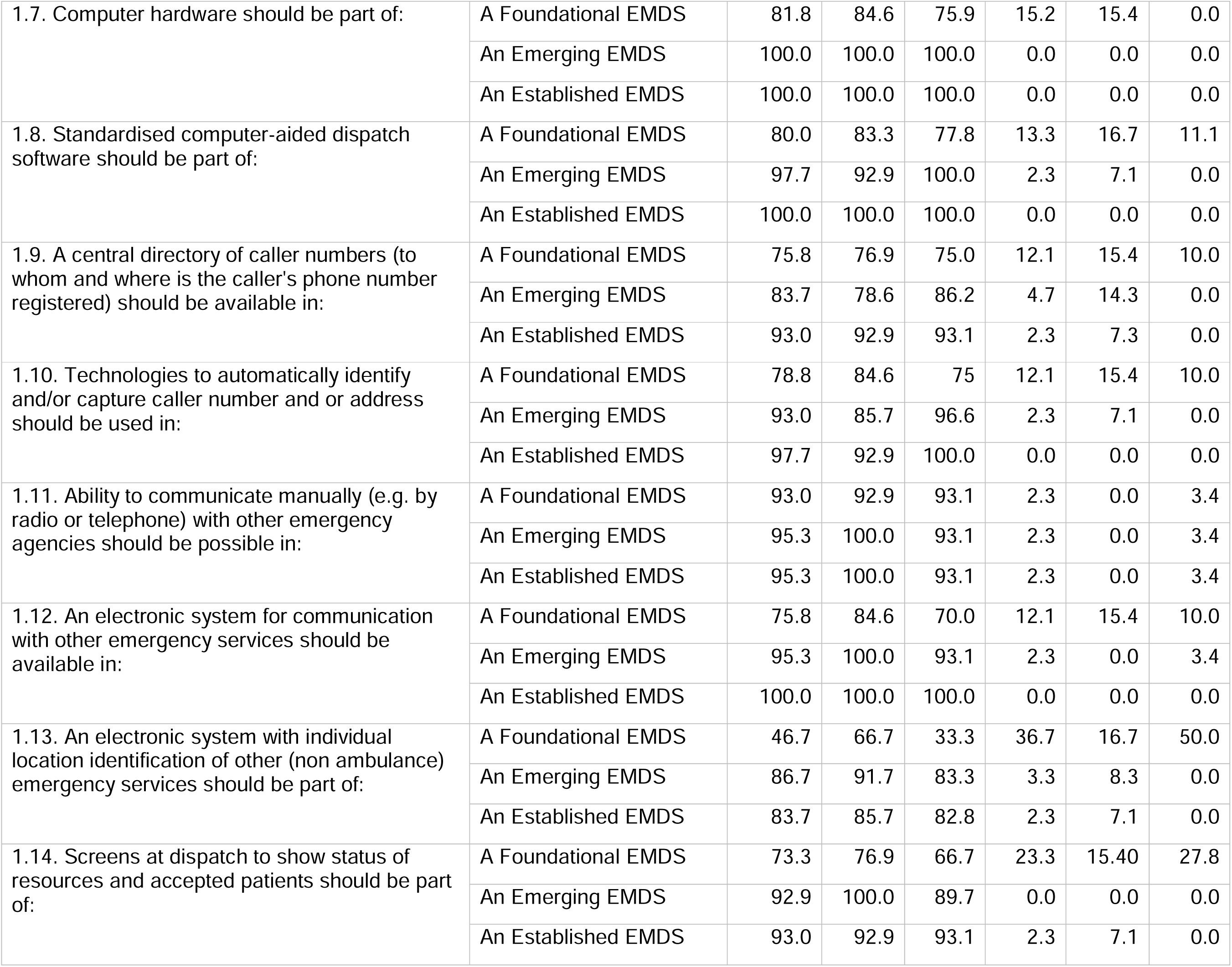
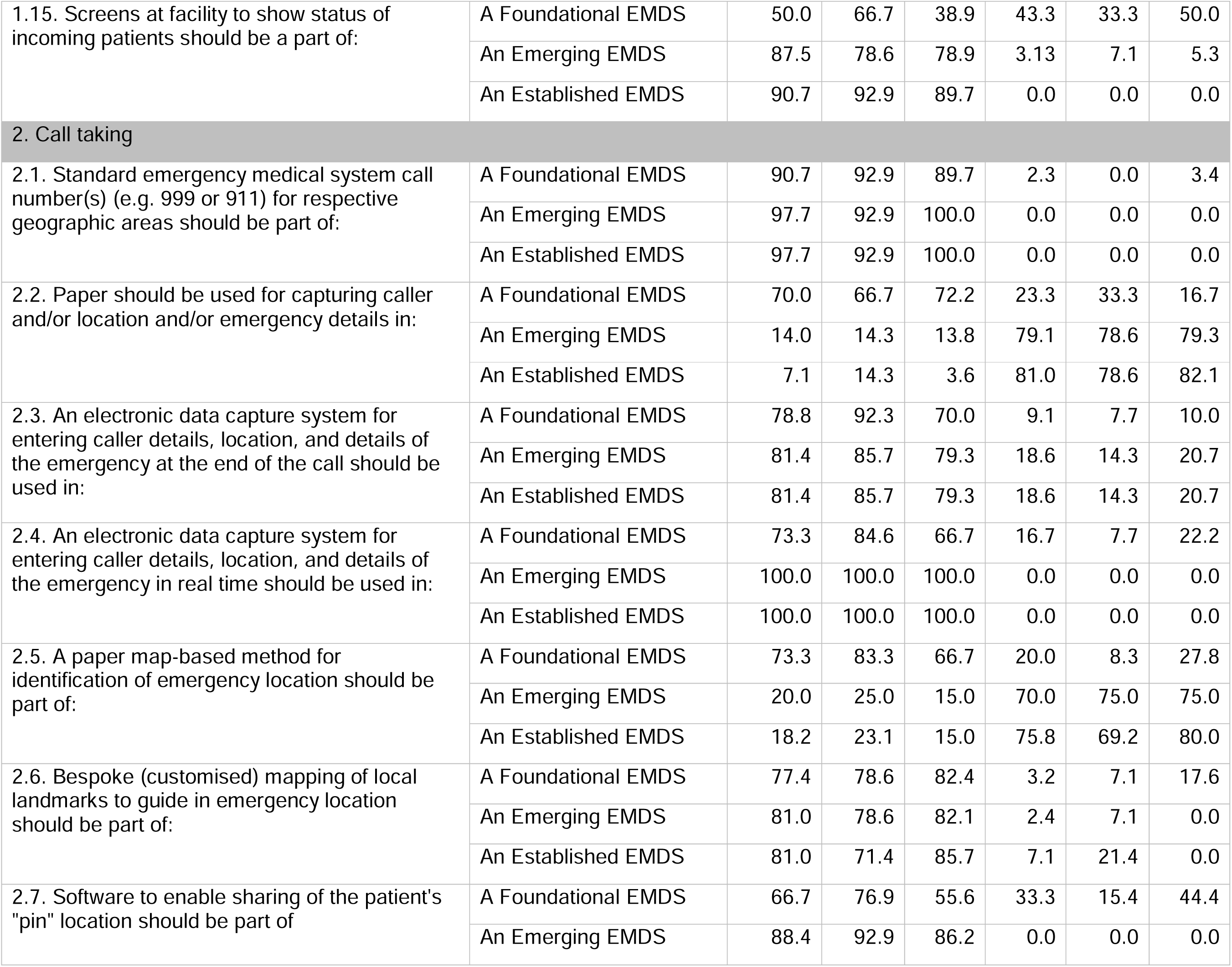
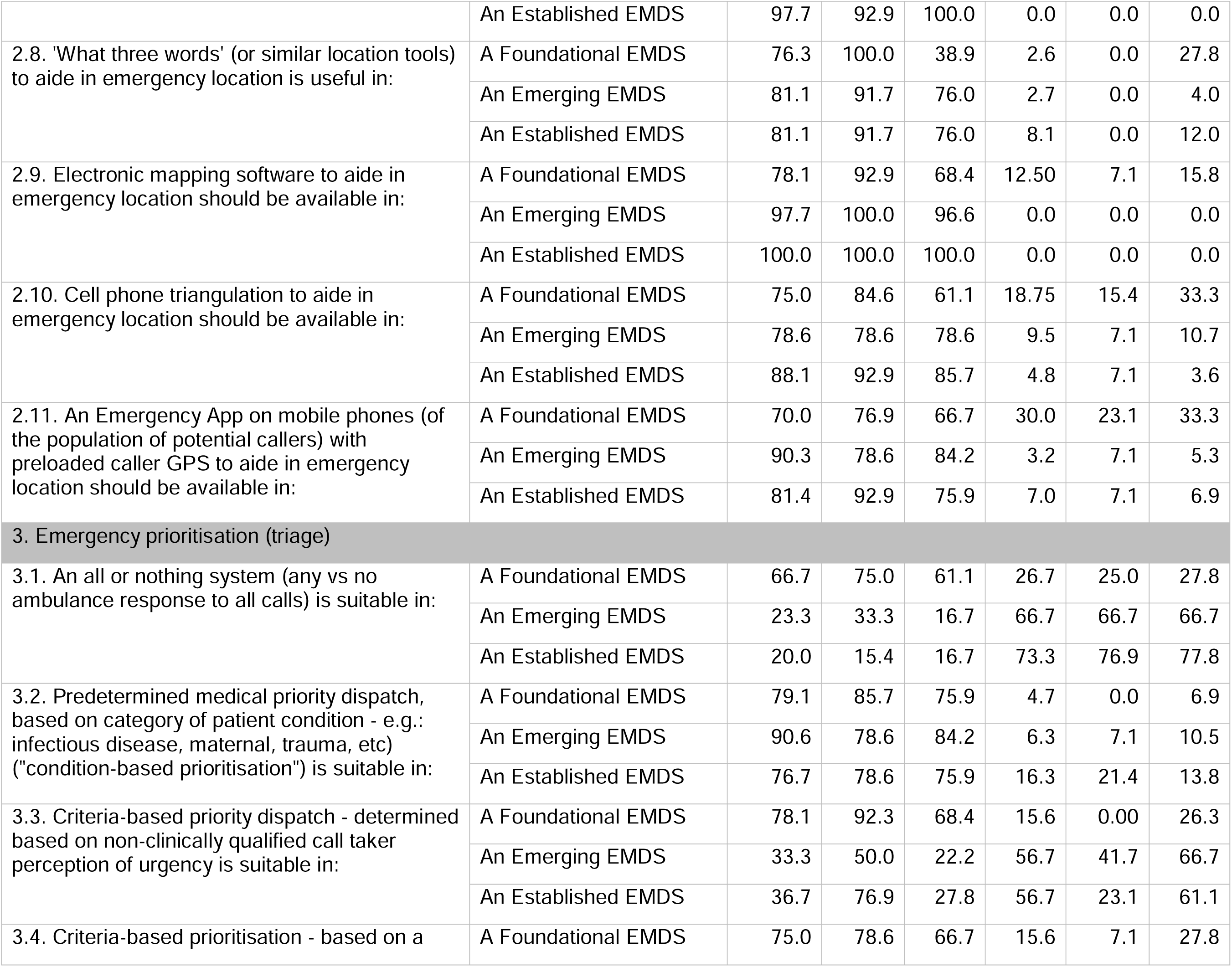
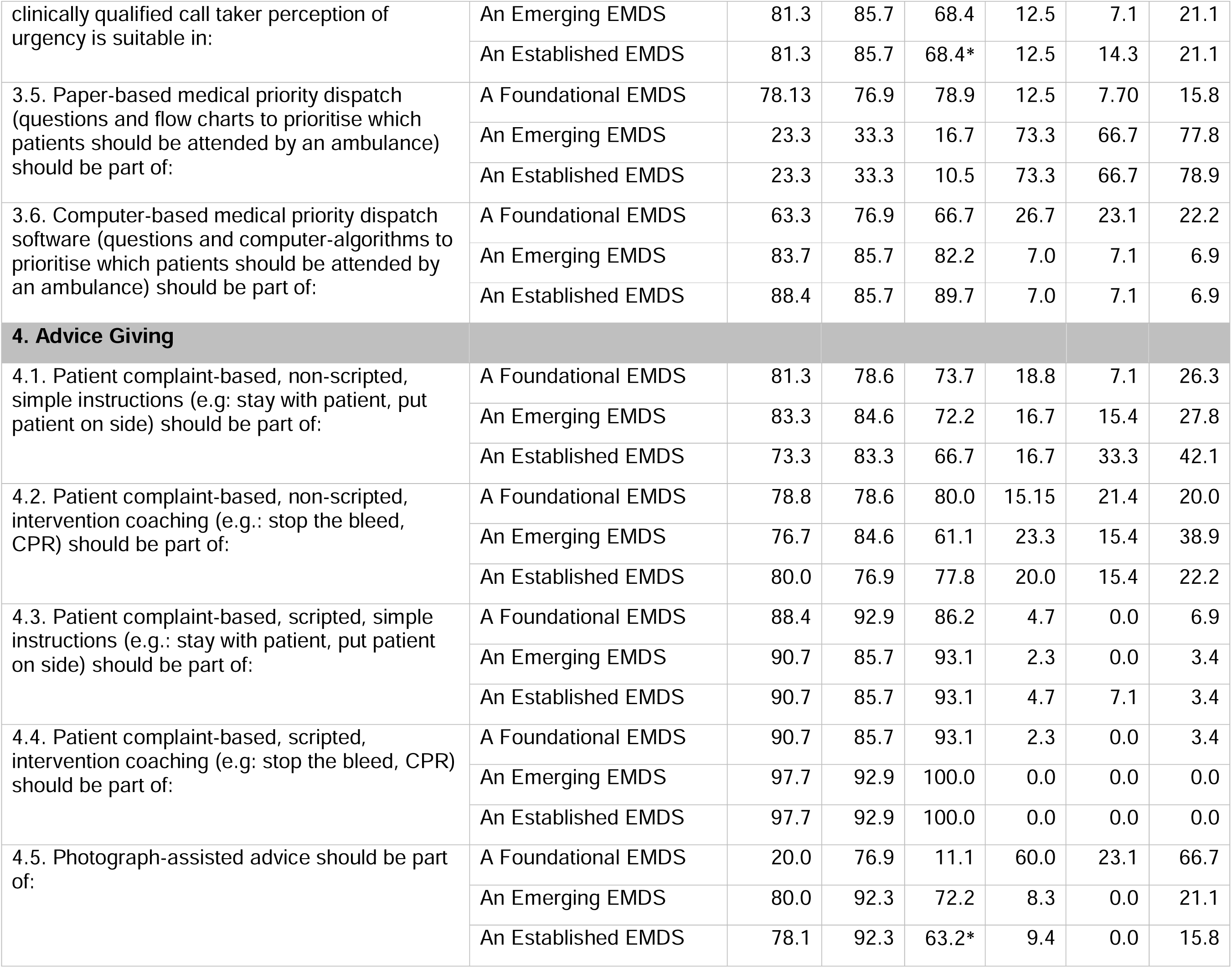
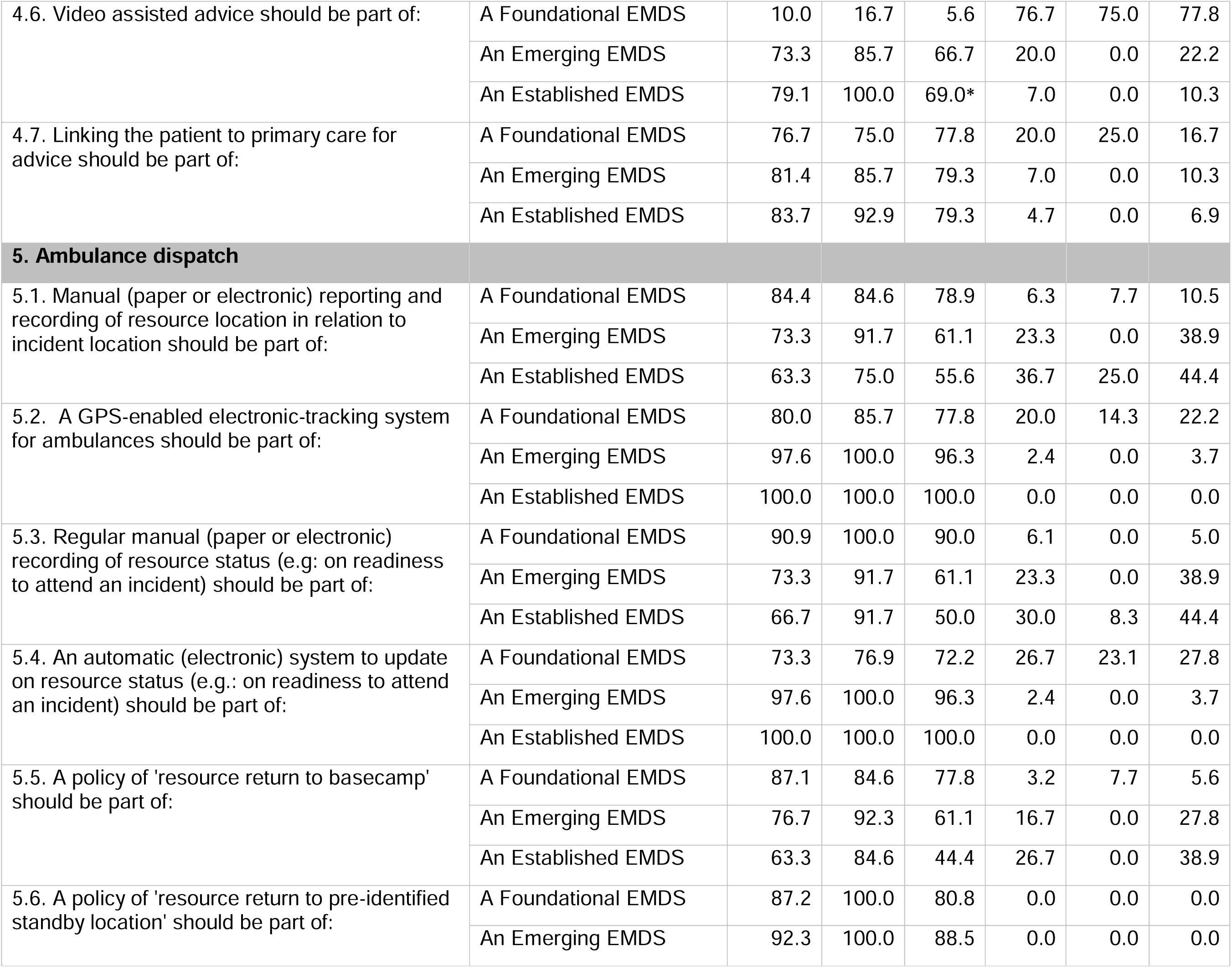
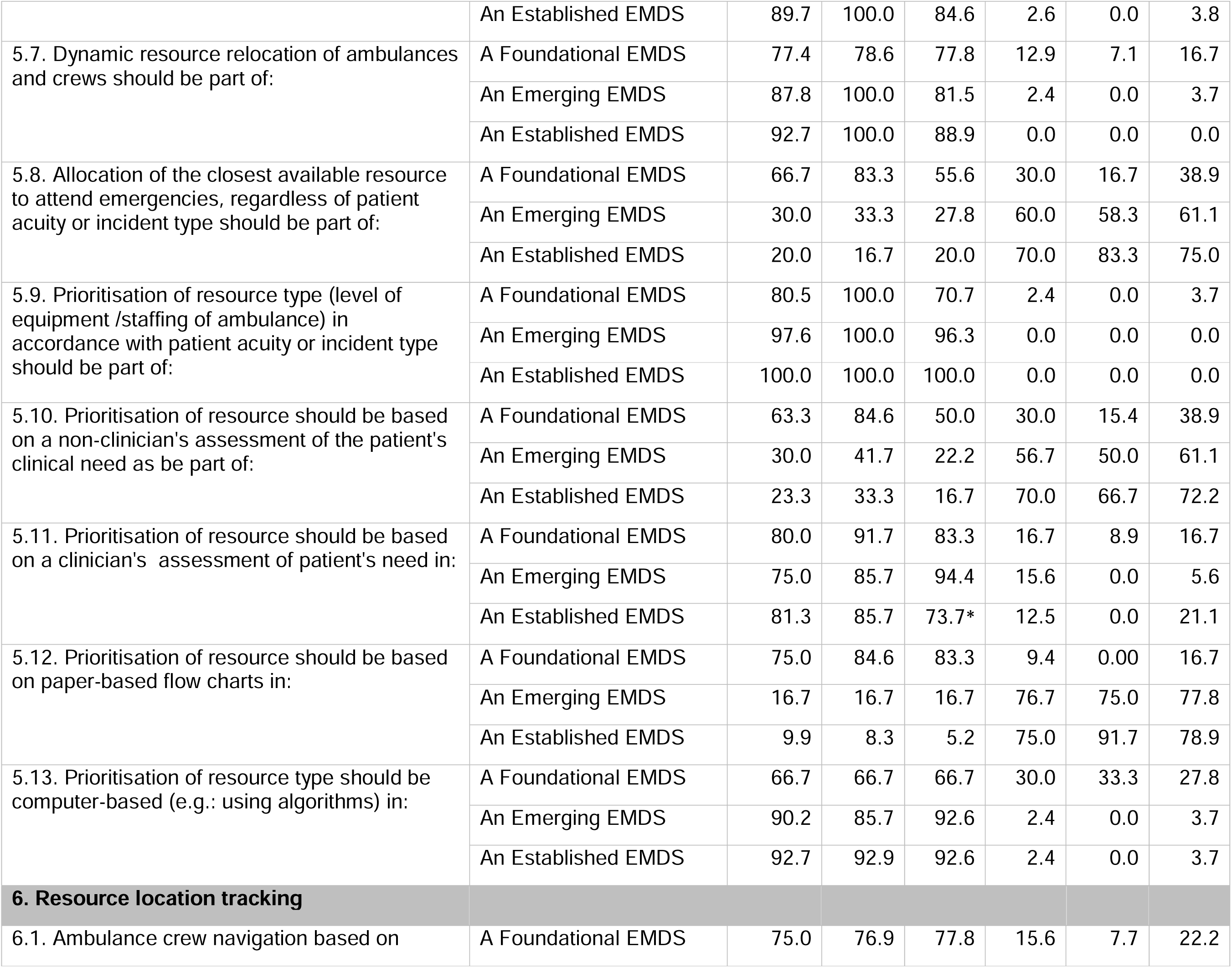
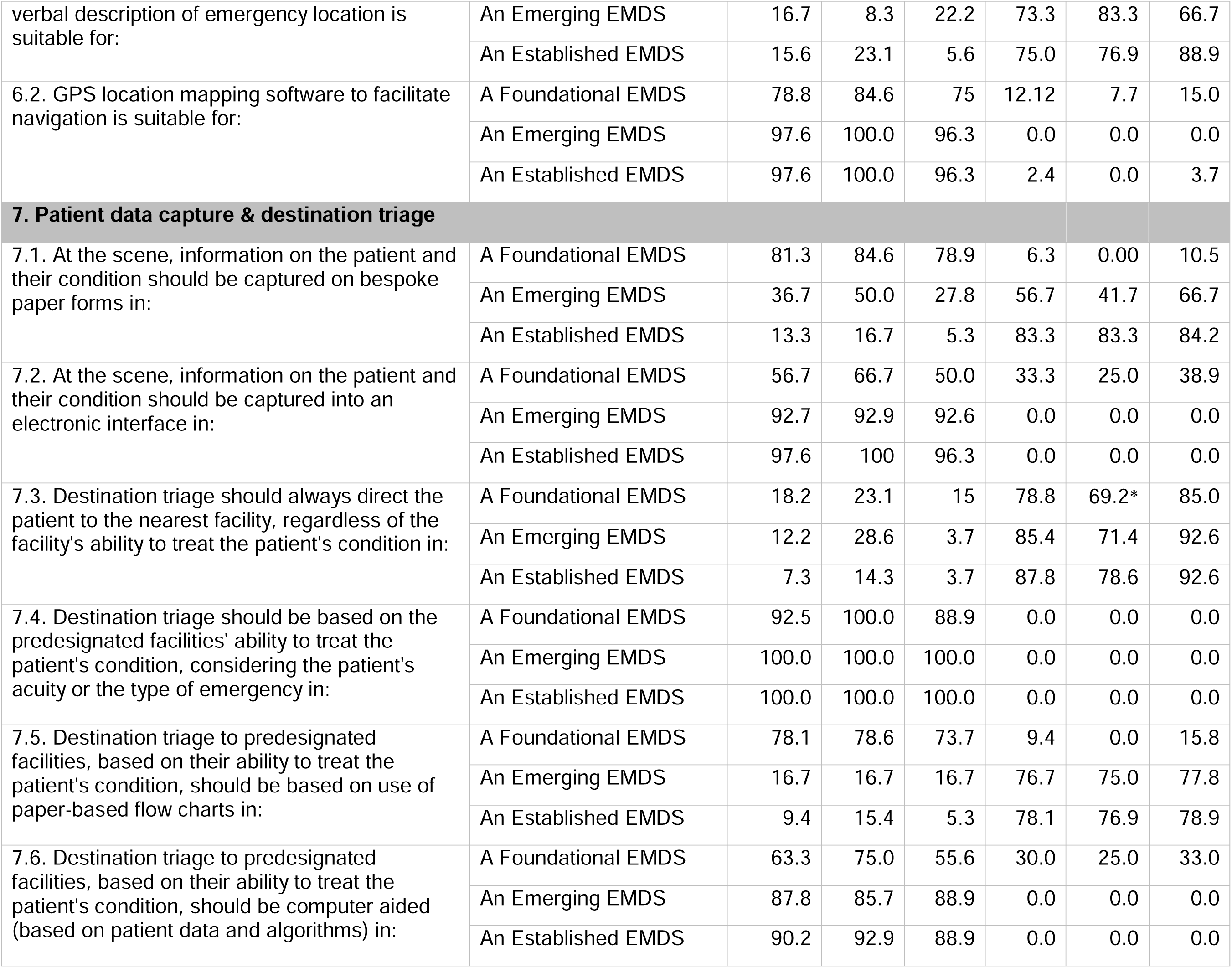
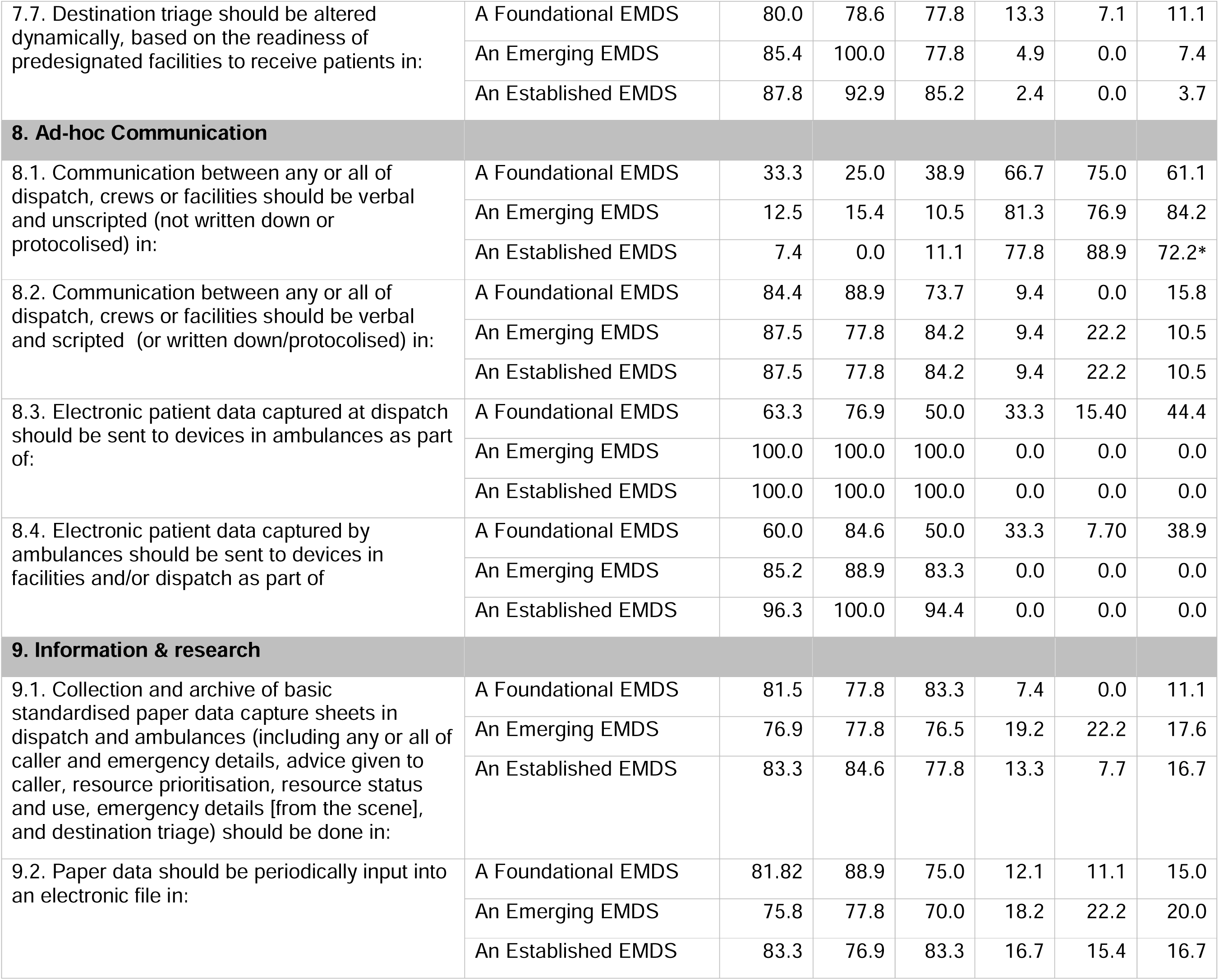
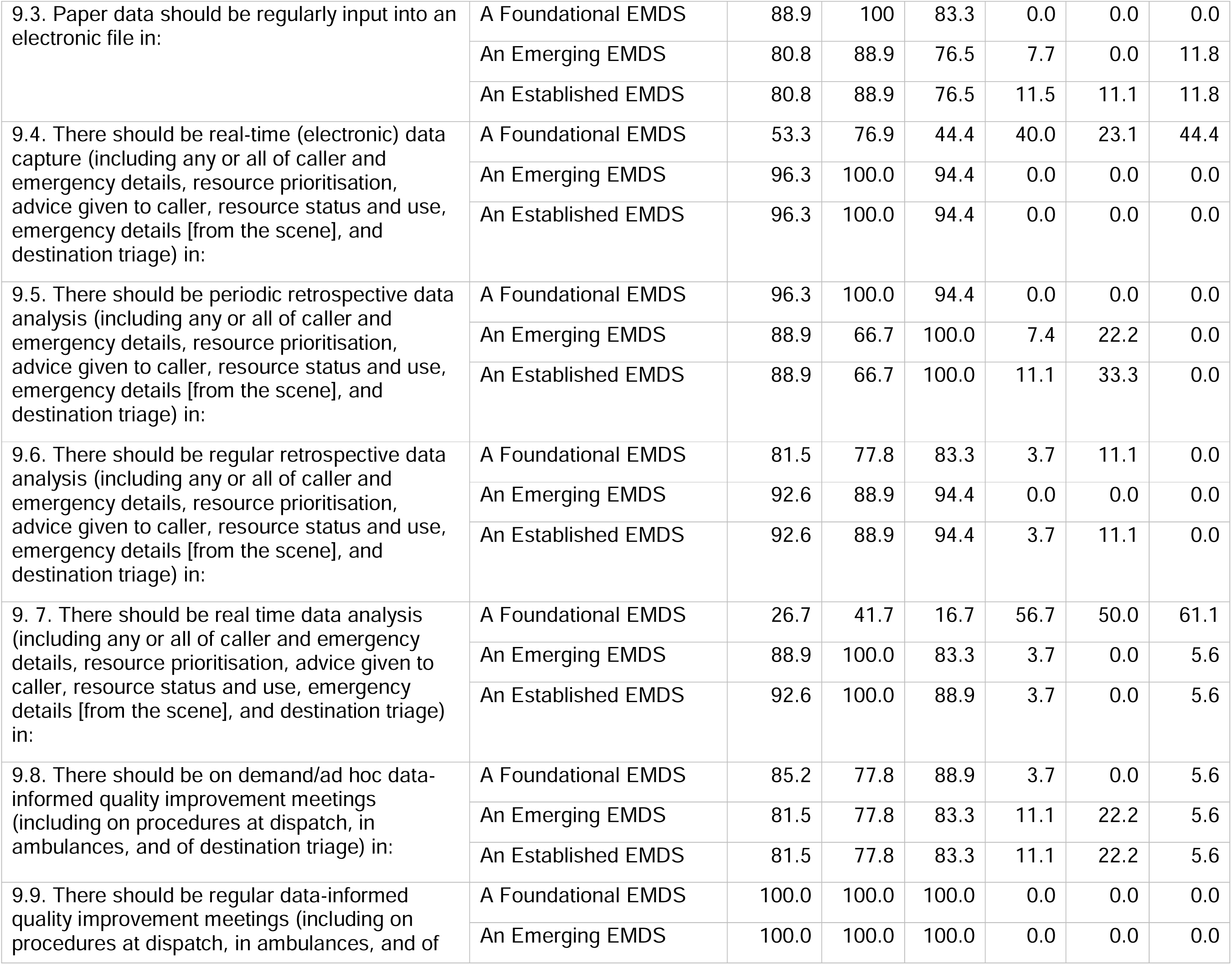
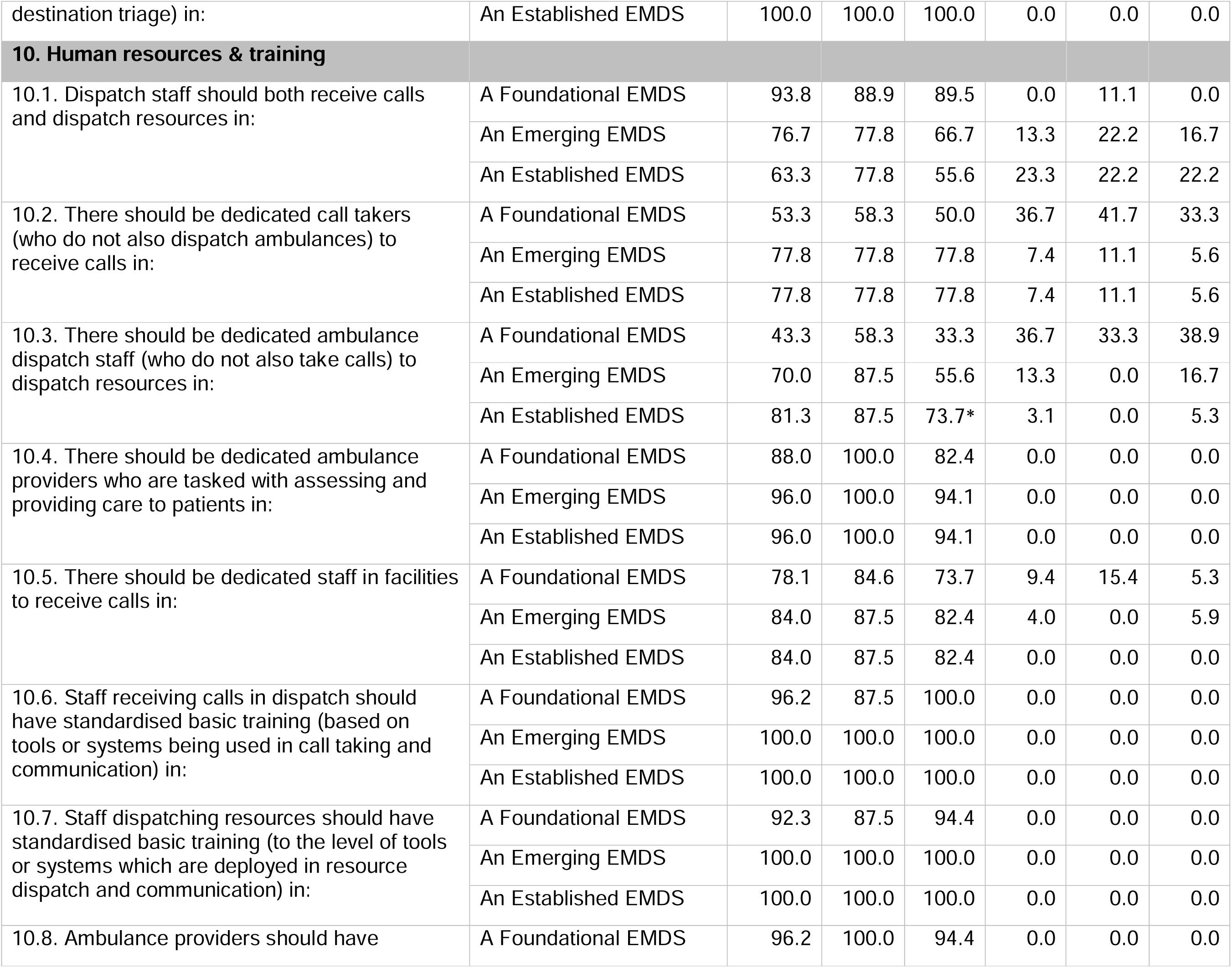
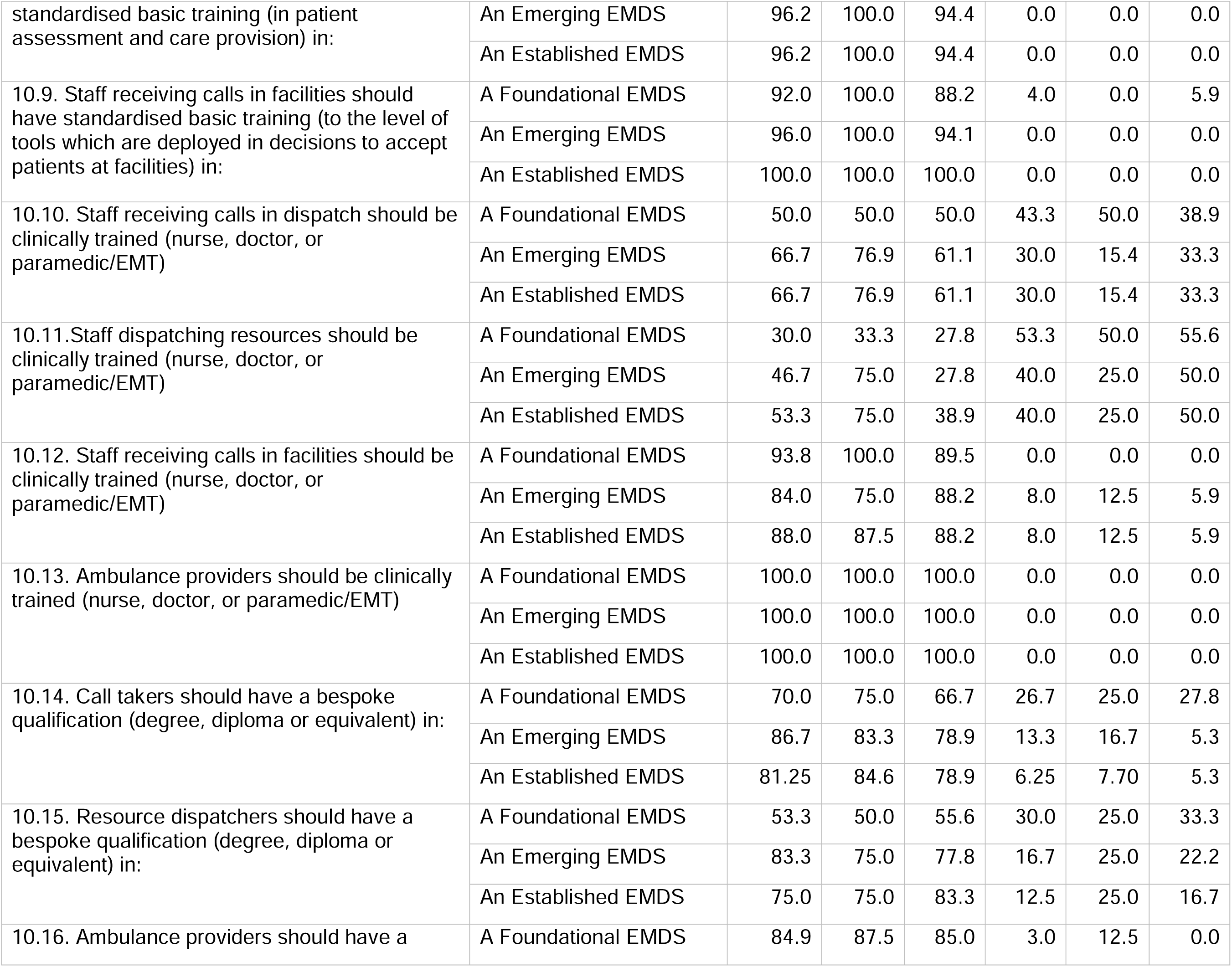
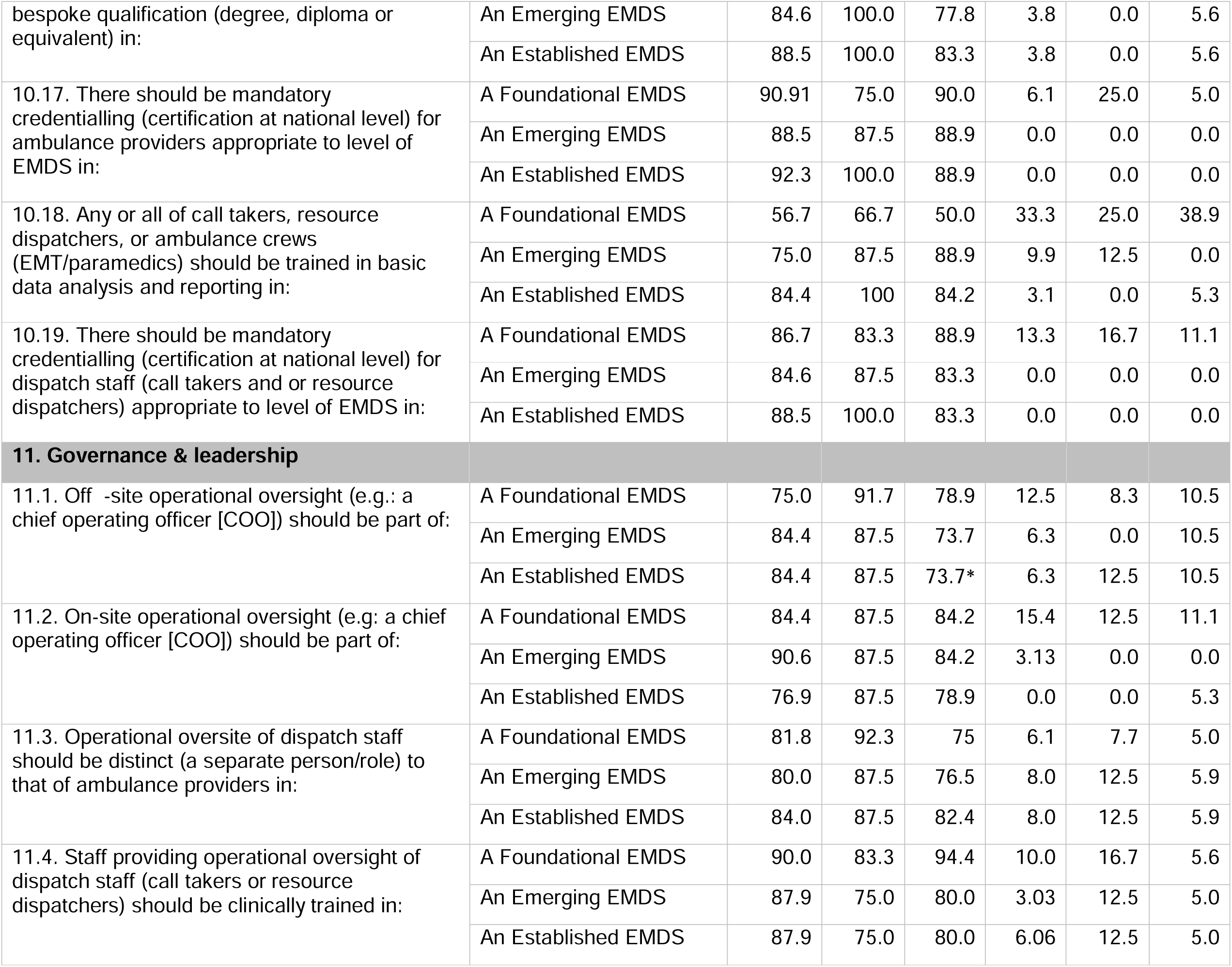
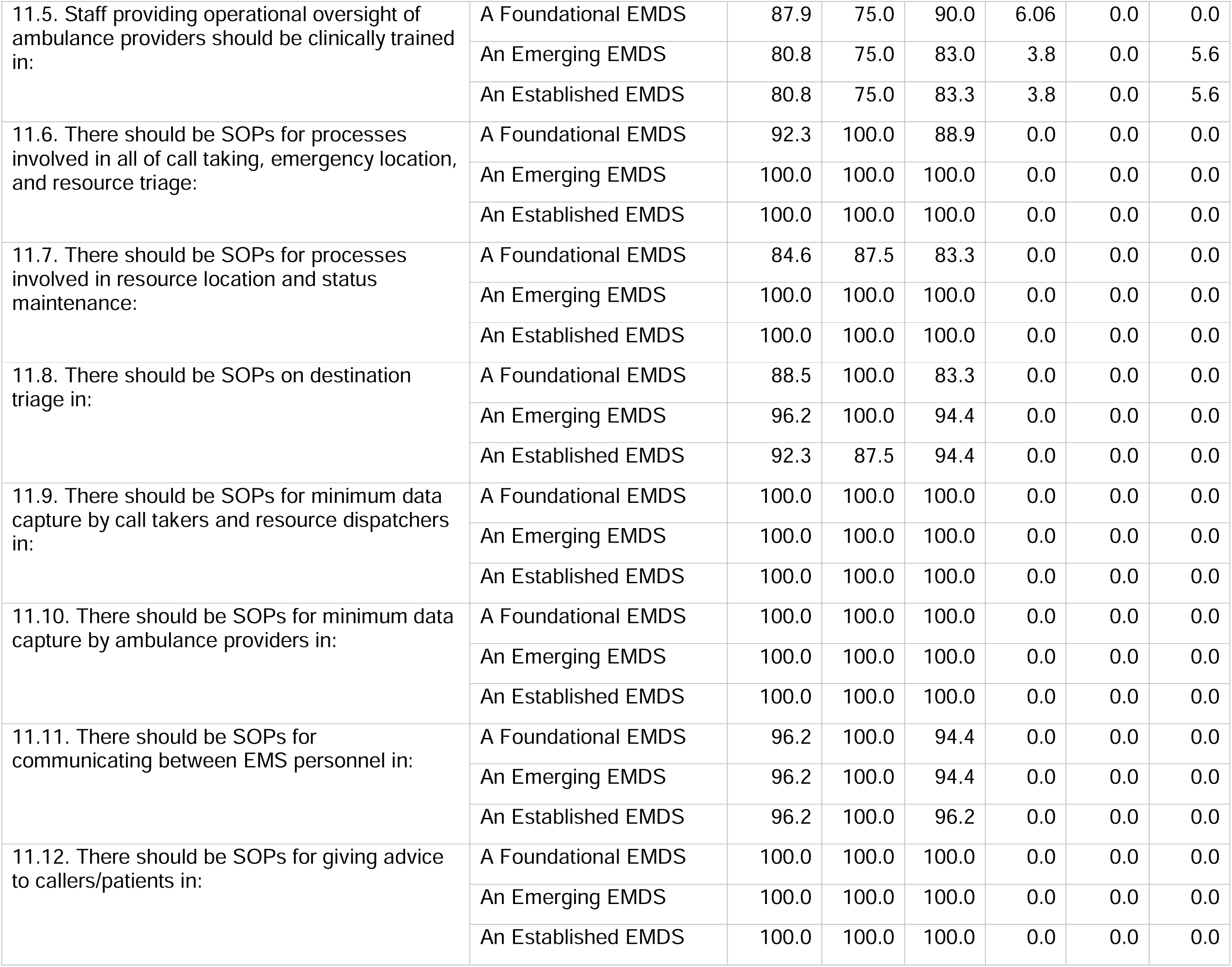
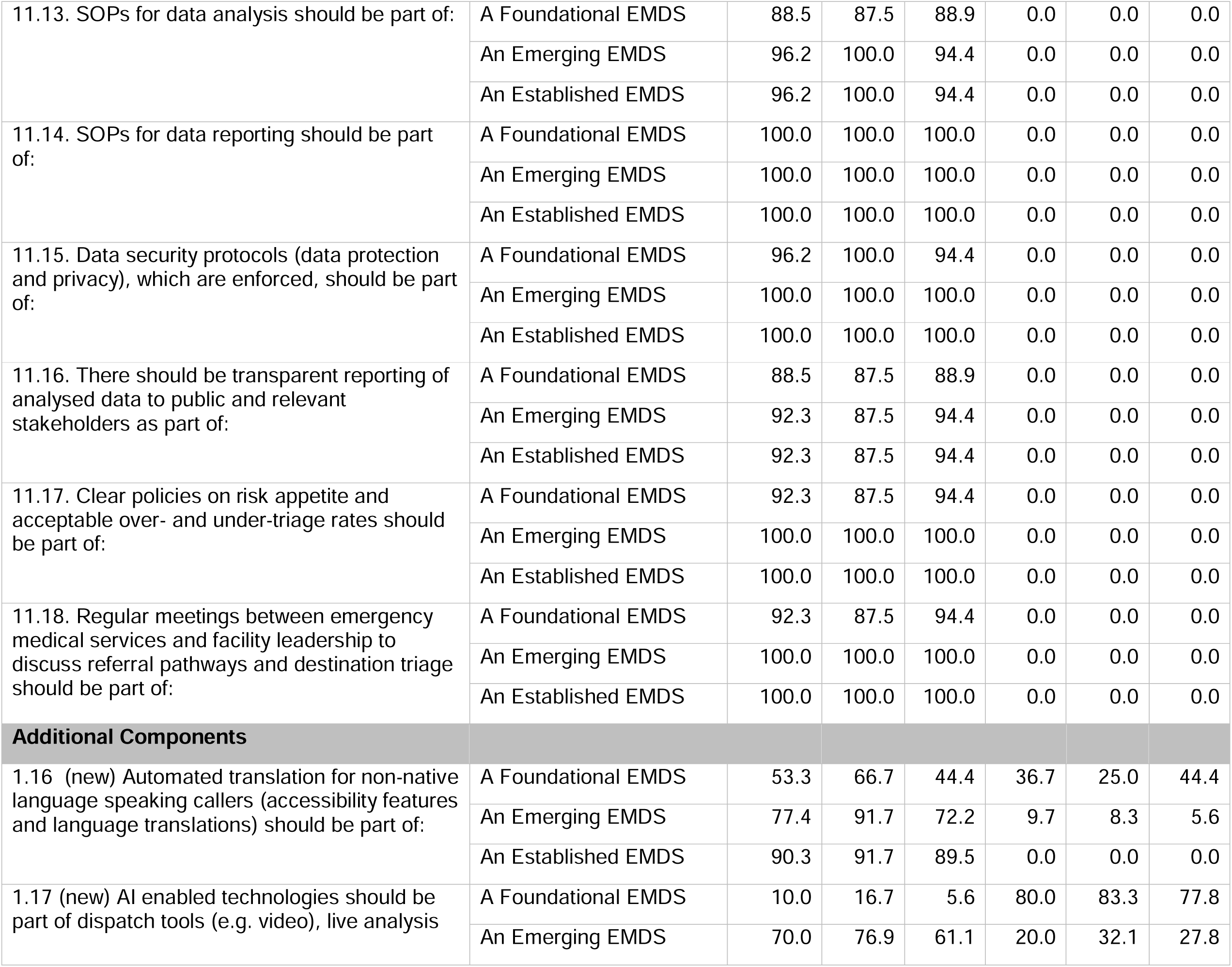
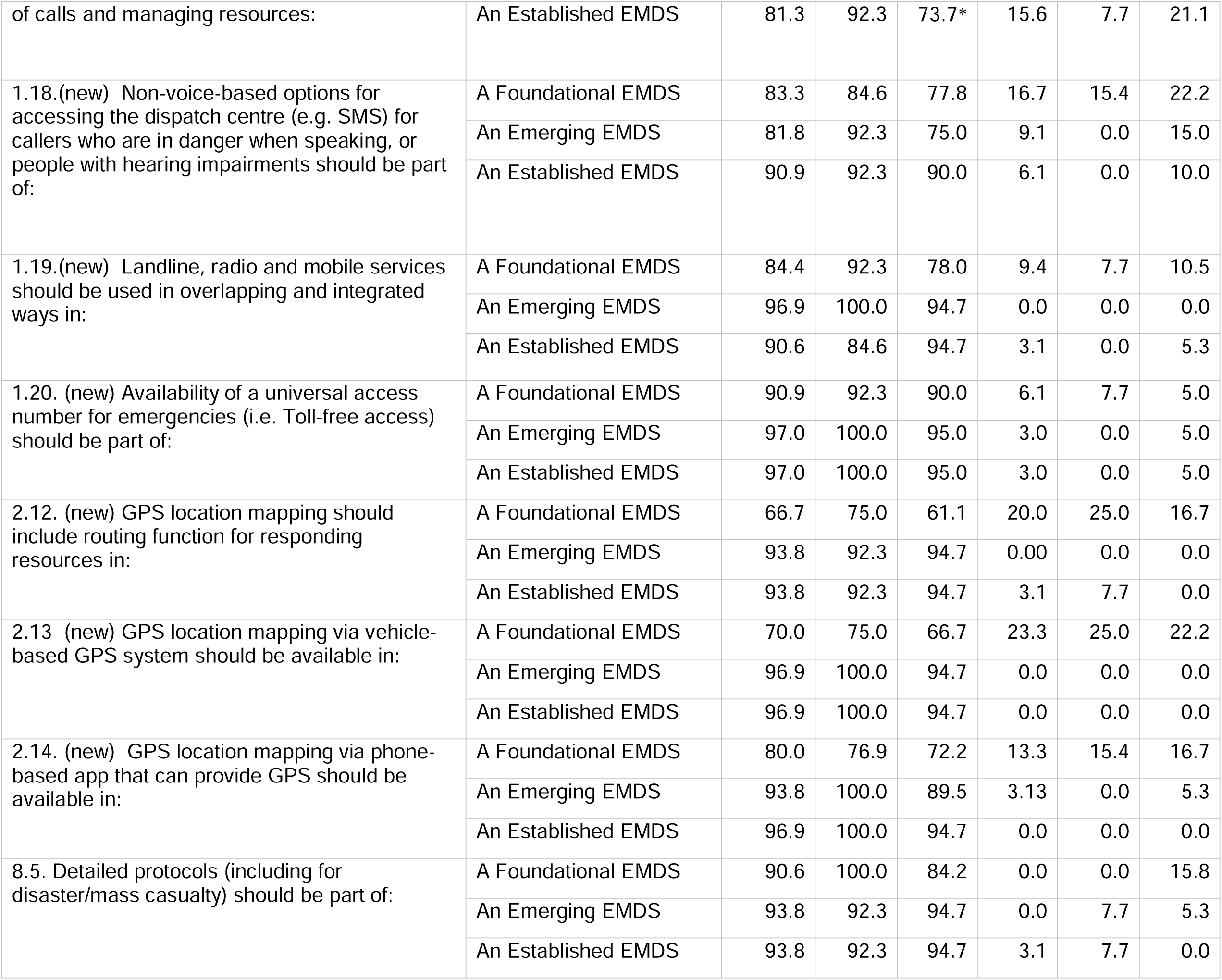
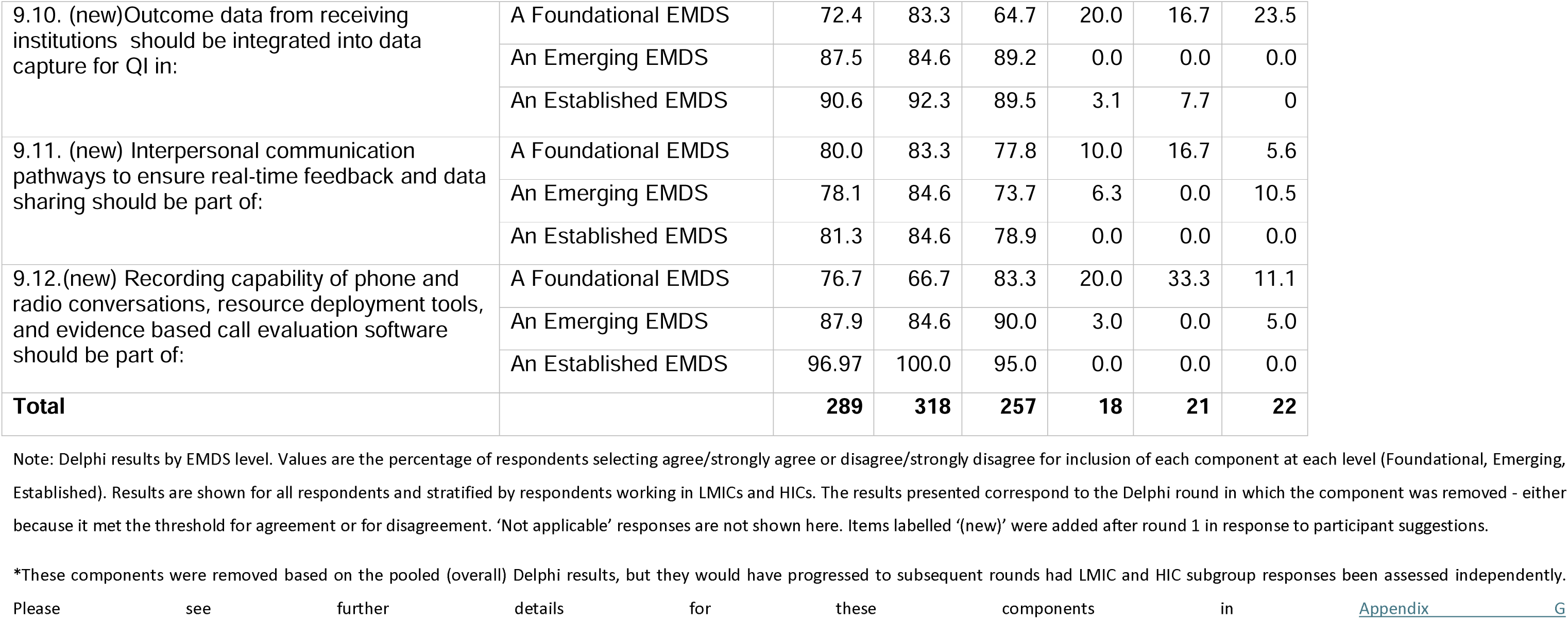

## Appendix E: Components agreed for inclusion by participants from one but not another income strata

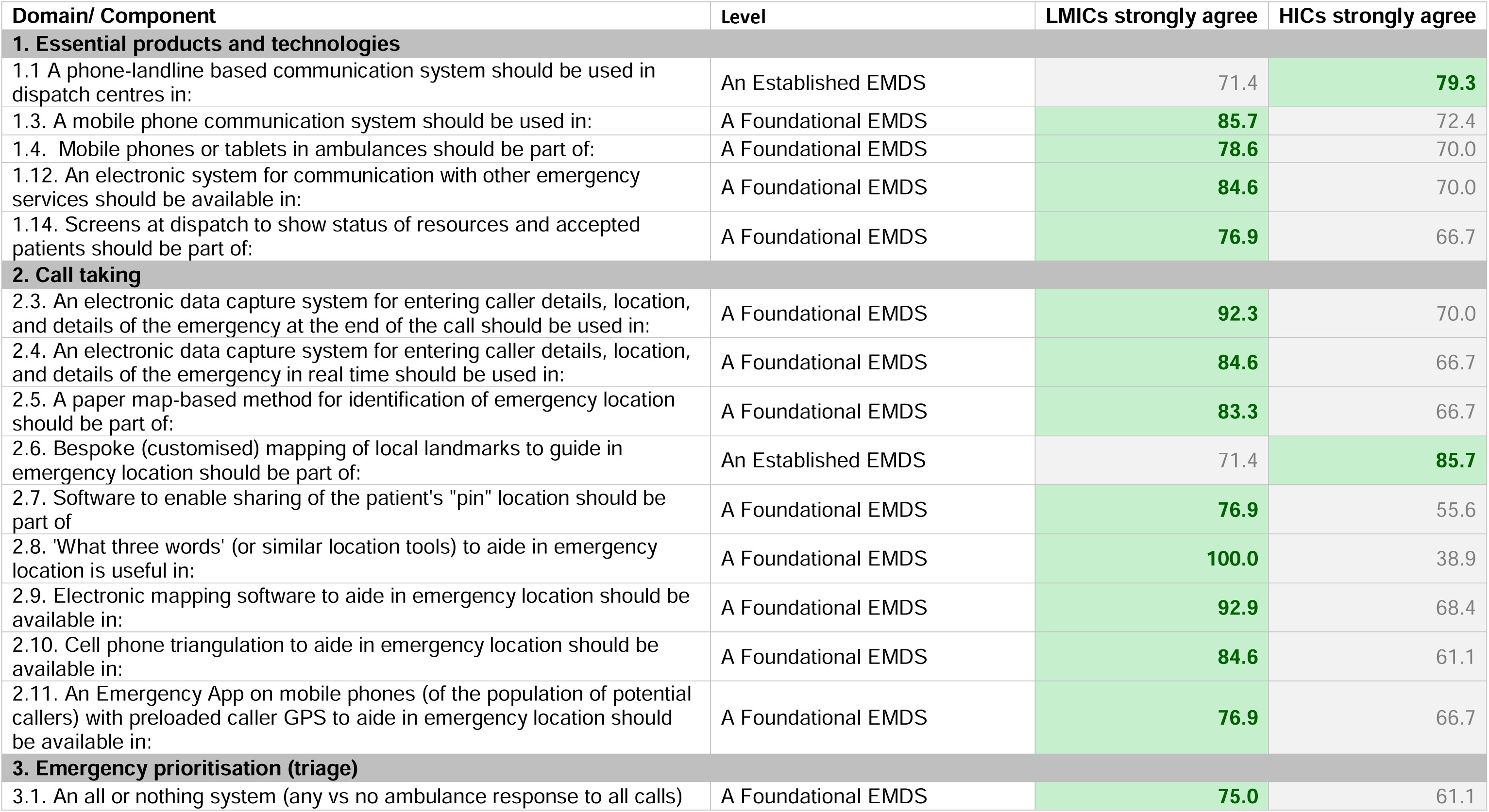
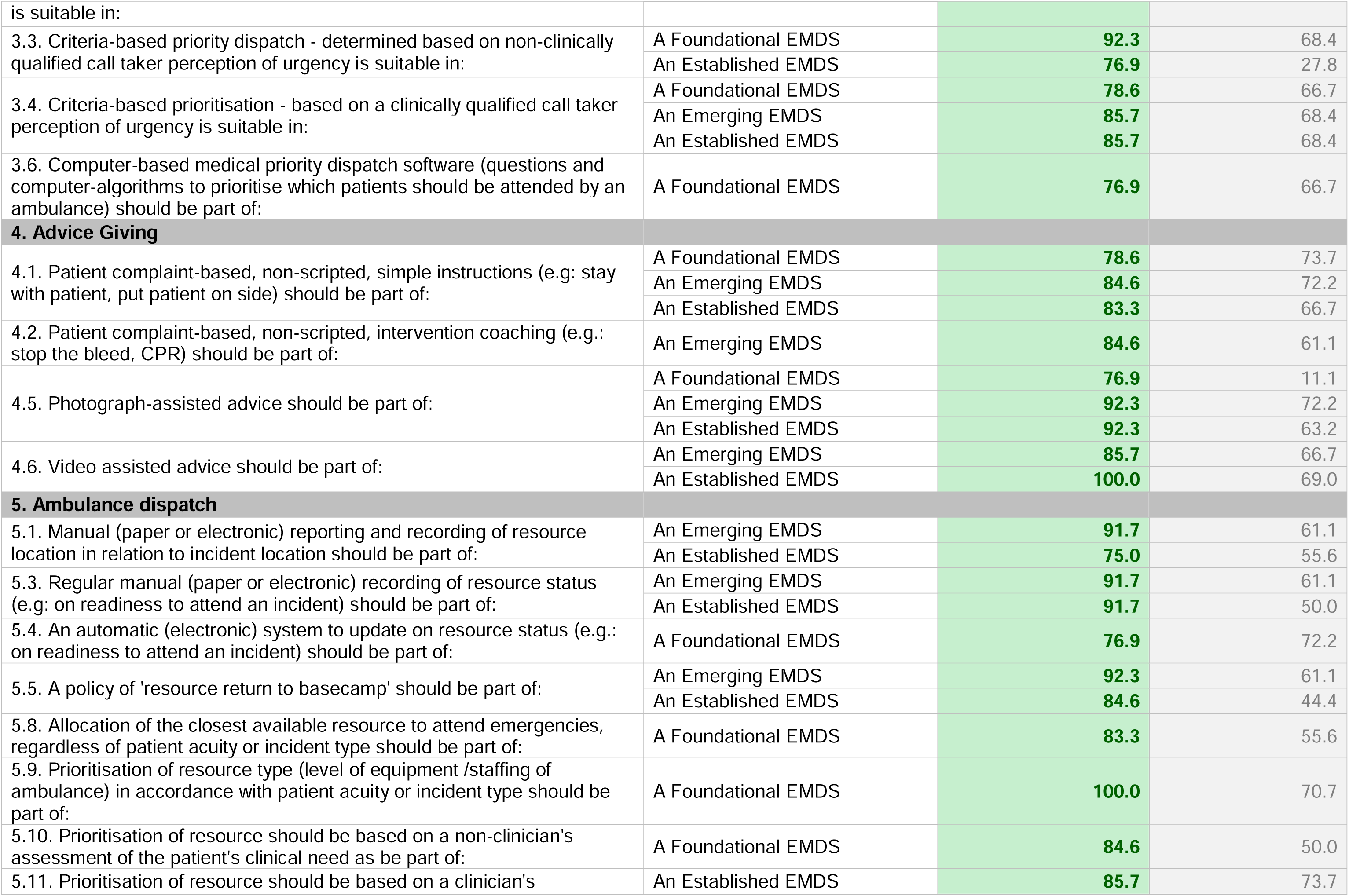
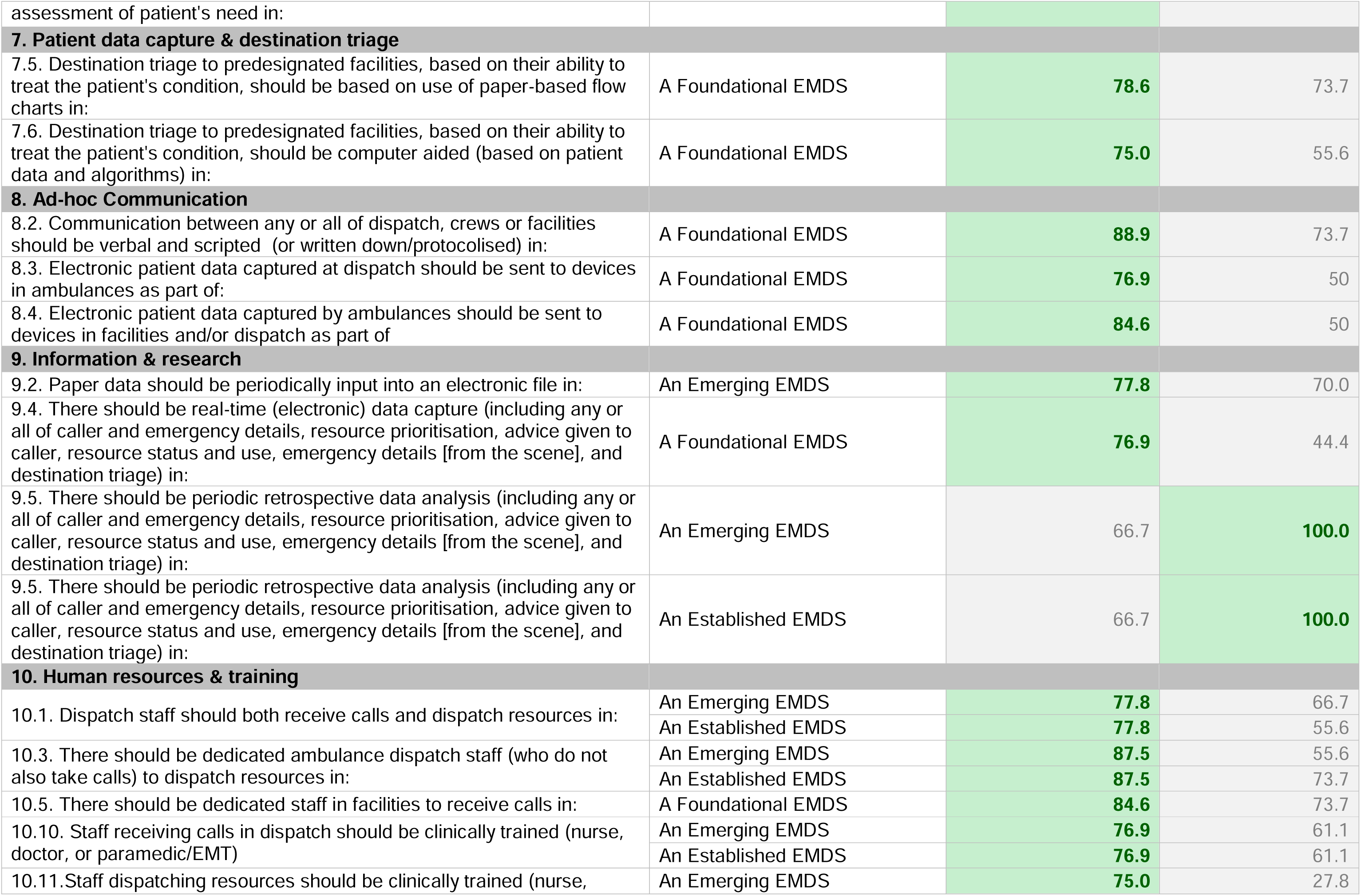
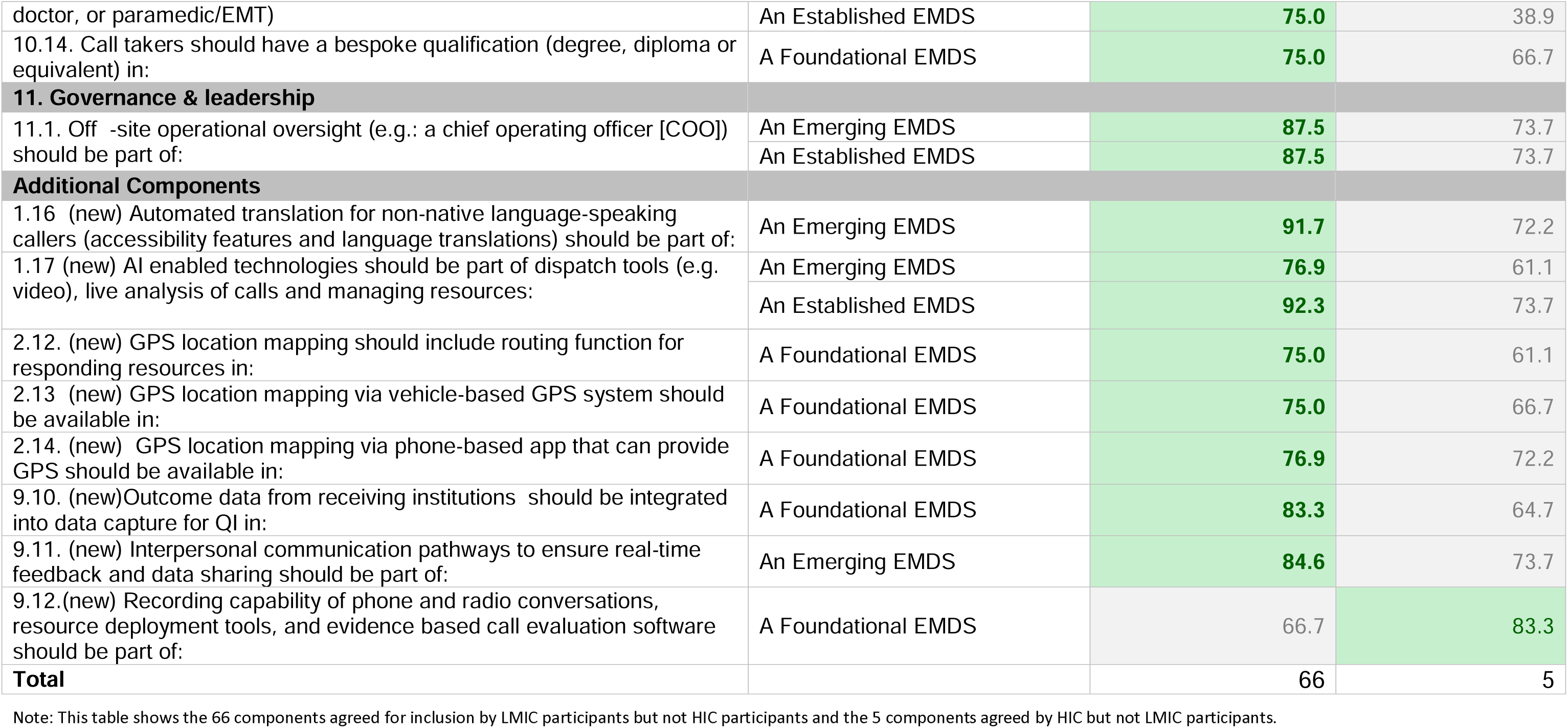

## Appendix F: Component Recommendations Based on IncomelzlDisaggregated Delphi Results

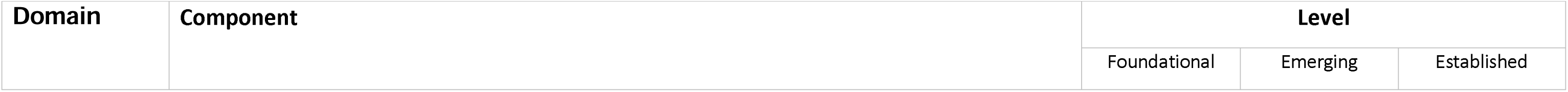
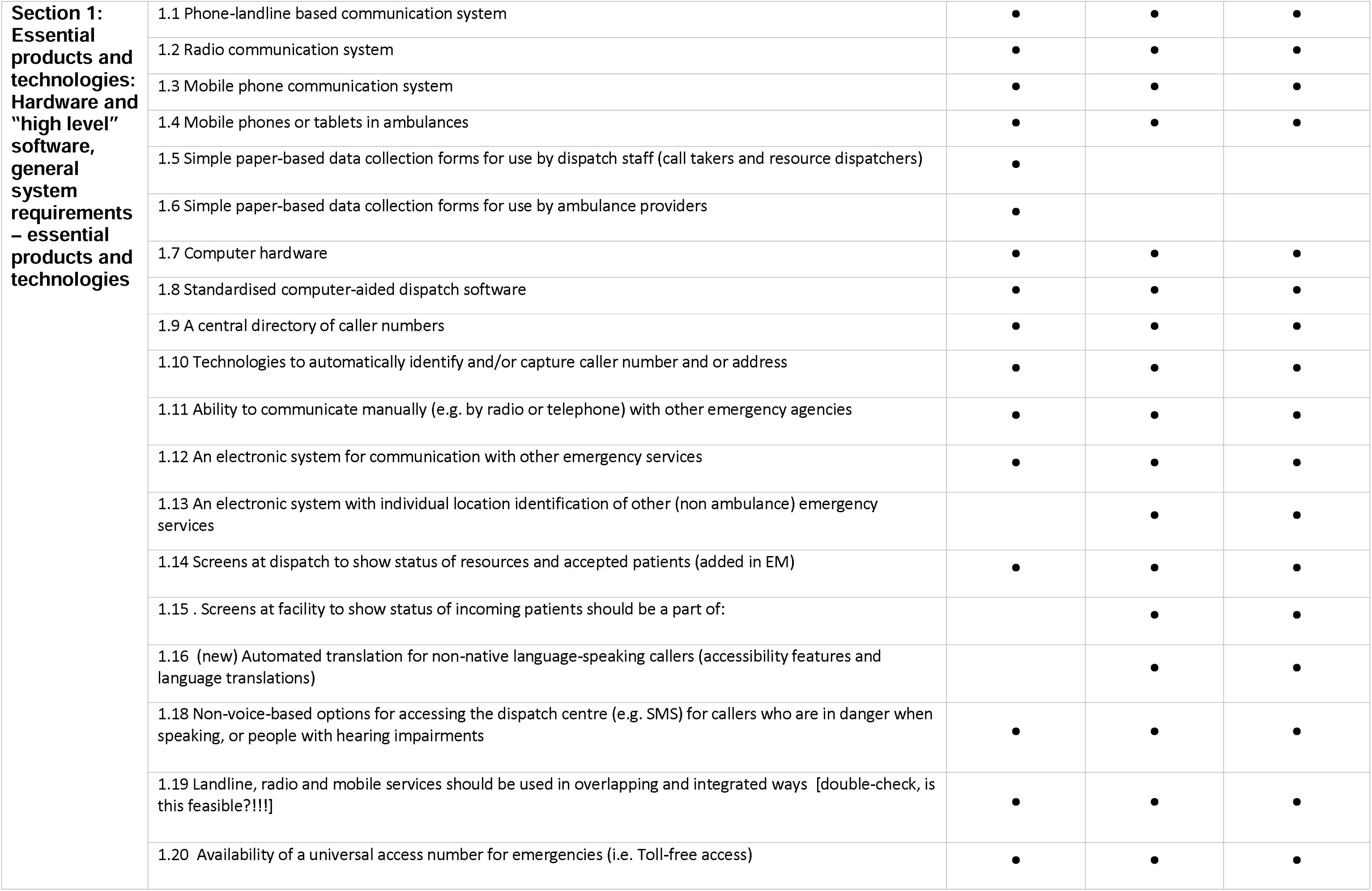
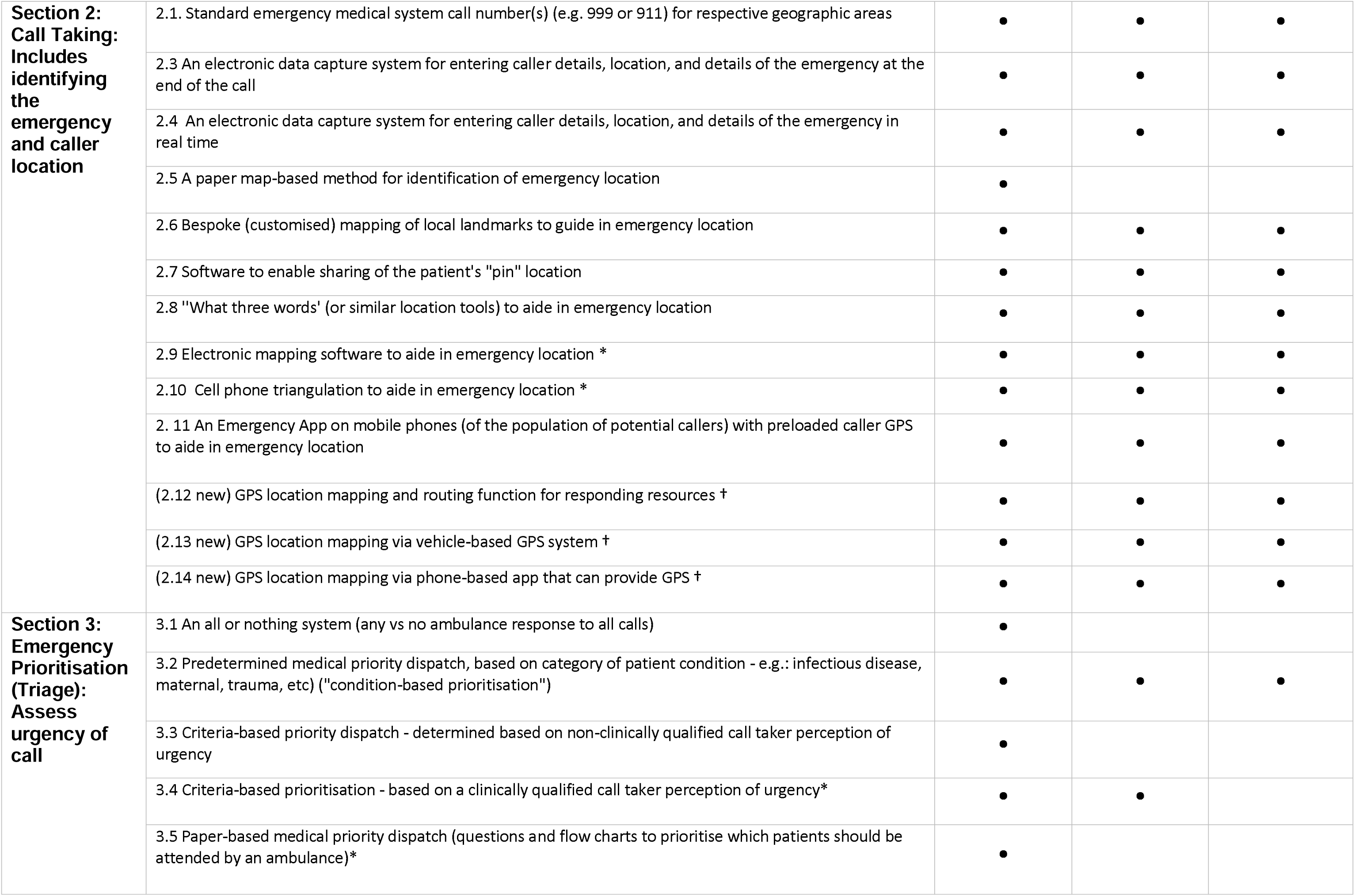
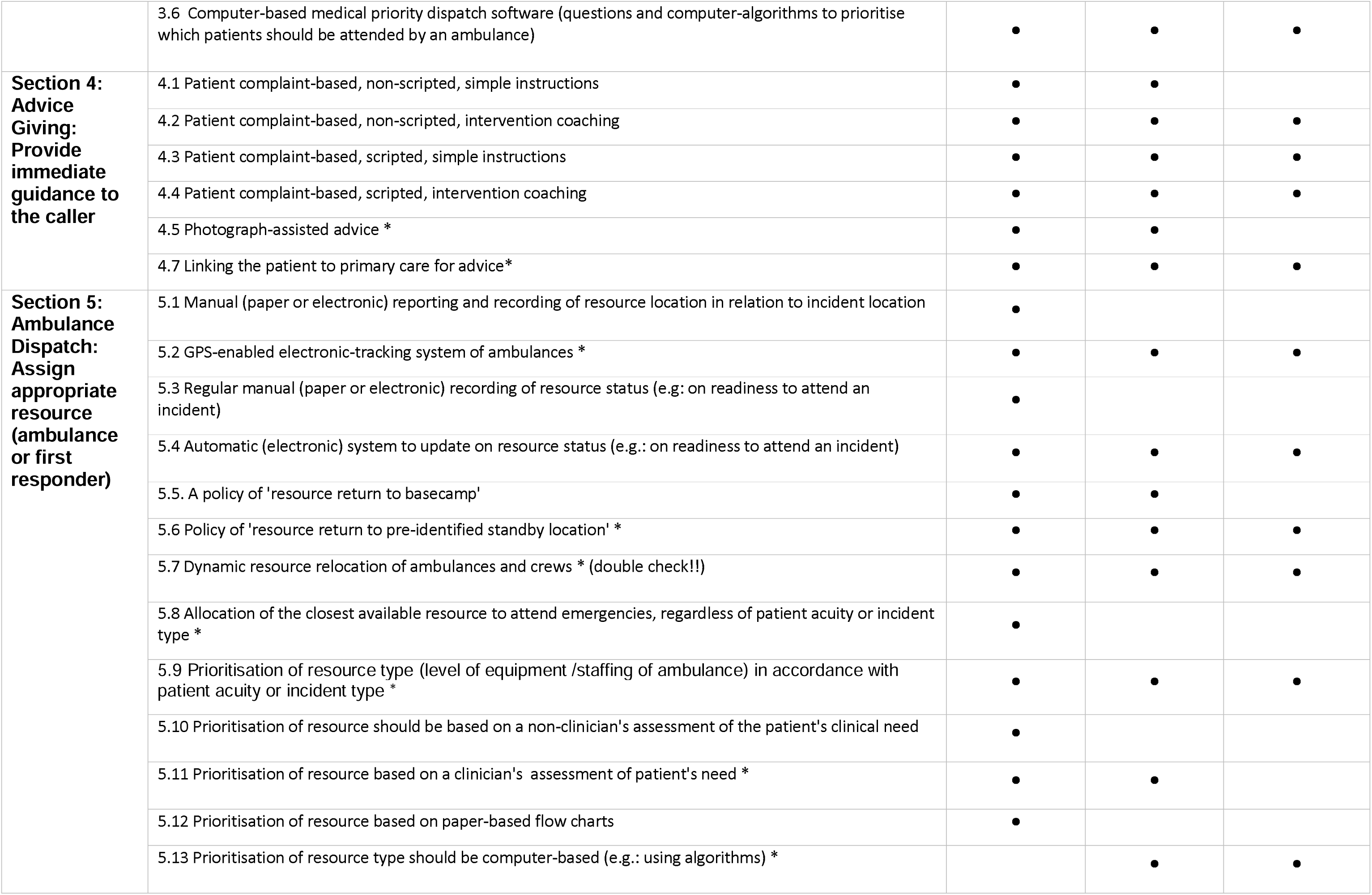
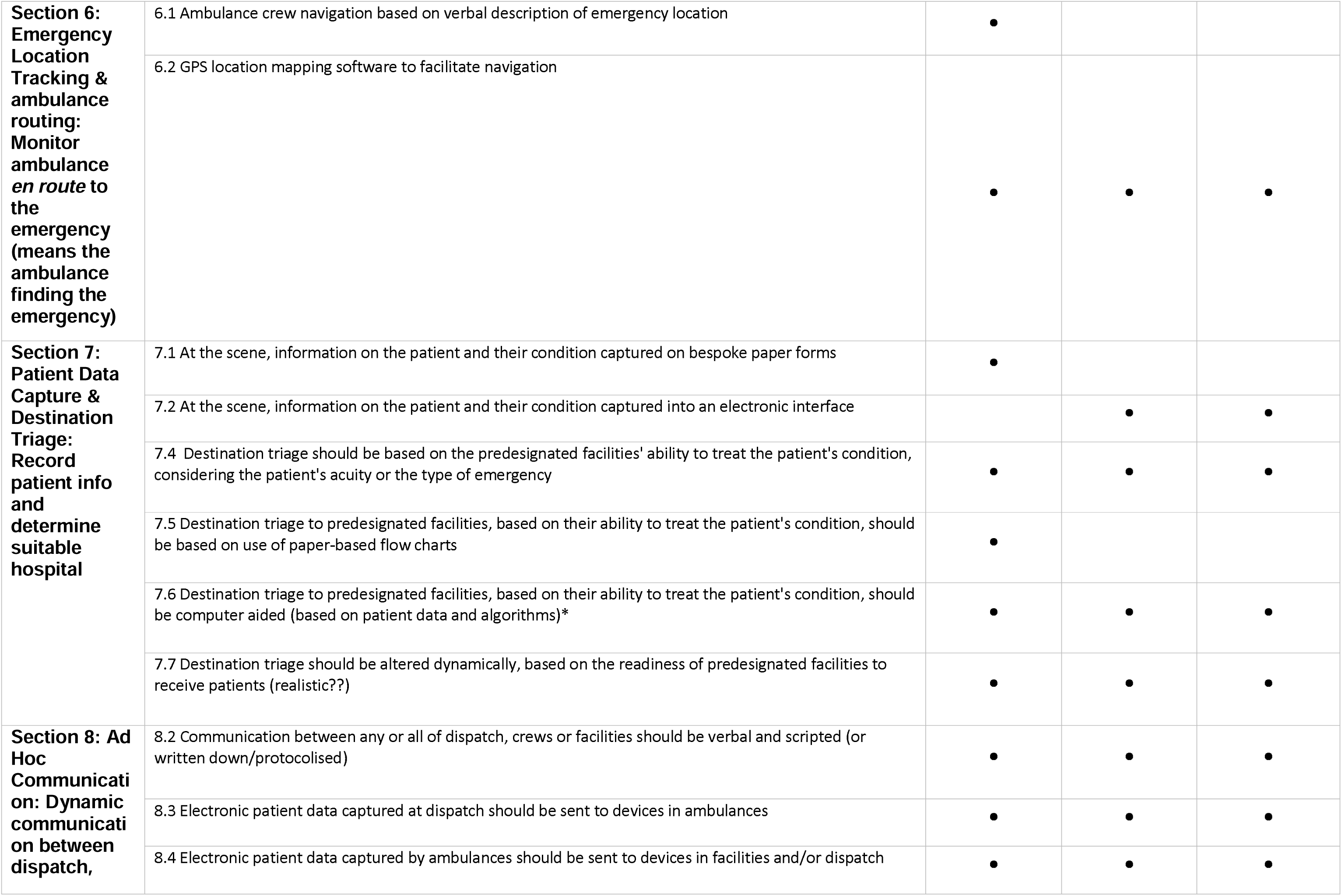
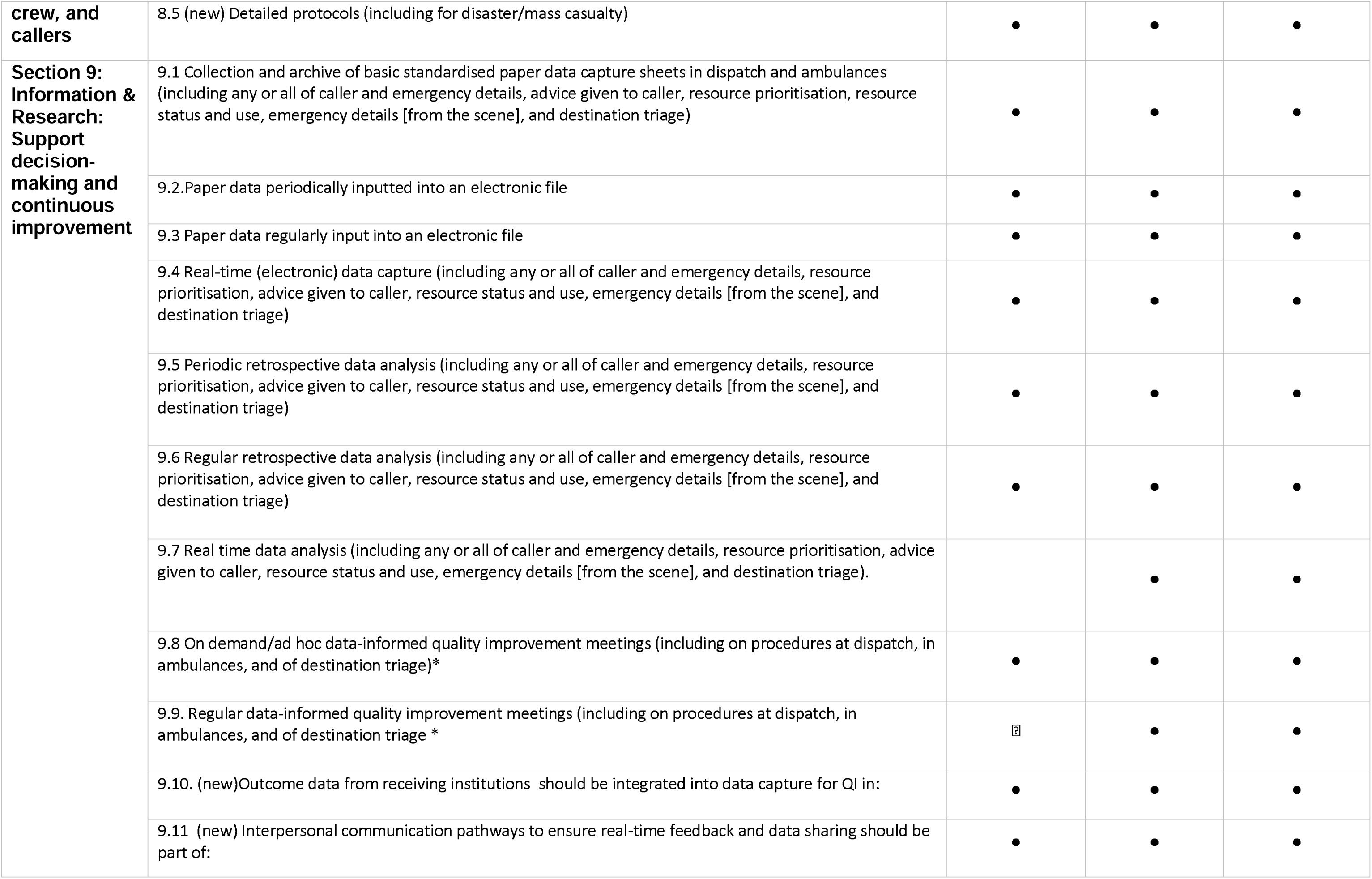
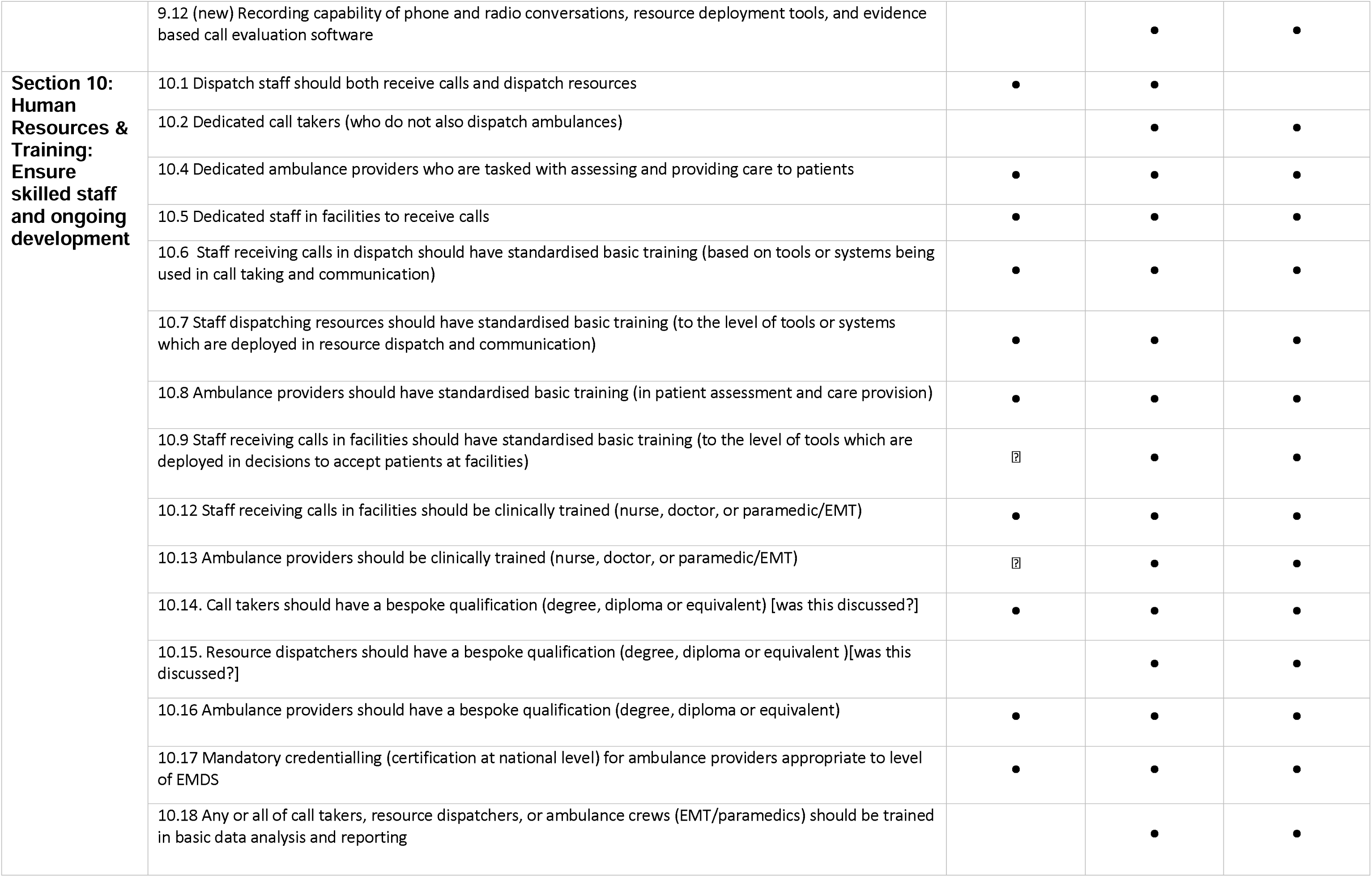
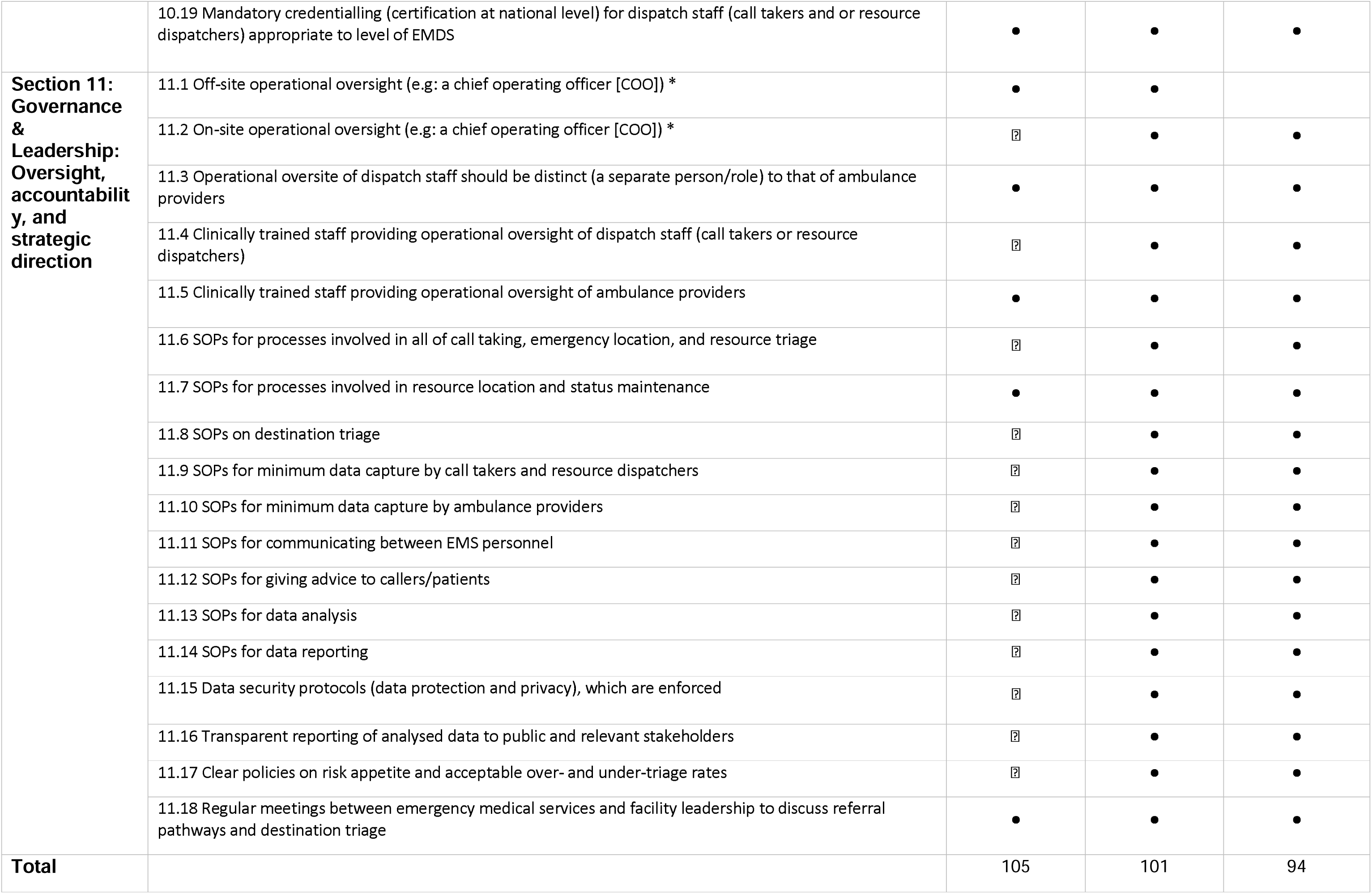
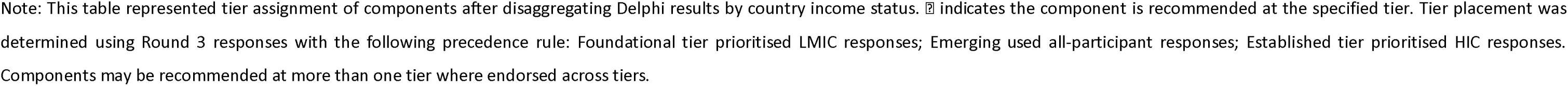

## Appendix G: Components “Erroneously” Removed from next round of Delphi for individual income strata analysis

| Domain | Component | Level Removed |  |  |
| --- | --- | --- | --- | --- |
|  |  | Foundational | Emerging | Established |
| ROUND 1 |  |  |  |  |
| 4. Advice Giving | 4.6. Video assisted advice |  |  | ? |
| 8. Ad-hoc Communication | 8.1. Communication between any or all of dispatch, crews or facilities to be verbal and unscripted (not written down or protocolised) |  |  | ? |
| ROUND 2 |  |  |  |  |
| 1. Essential products and technologies | 1.17 (new) AI enabled technologies to be part of dispatch tools (e.g. video), live analysis of calls and managing resources |  |  | ? |
| 3. Emergency prioritisation (triage) | 3.4. Criteria-based prioritisation - based on a clinically qualified call taker perception of urgency |  |  | ? |
| 4. Advice Giving | 4.5 Photograph-assisted advice |  |  | ? |
| 5. Ambulance dispatch | 5.11. Prioritisation of resource should be based on a clinician's assessment of patient's need |  |  | ? |
| 7. Patient data capture & destination triage | 7.3. Destination triage should always direct the patient to the nearest facility, regardless of the facility's ability to treat the patient's condition in: | ? |  |  |
| 10. Human resources & training | 10.3. There should be dedicated ambulance dispatch staff (who do not also take calls) to dispatch resources |  |  | ? |
| 11. Governance & leadership | 11.1 Off -site operational oversight (e.g.: a chief operating officer [COO]) should be part of: |  |  | ? |

Table shows: Components affected by retrospective (after final round) analysis of pooled vs subgroup Delphi progression. This table lists items that were removed during the pooled Delphi process but would have progressed under subgroup-specific scoring.

